# Differential Associations Between Social Determinants of Health and the Initiation of Medications for Opioid Use Disorder Across Care Settings

**DOI:** 10.1101/2025.06.23.25330147

**Authors:** Zhen Luo, Mackenzie Hofford, Ruochong Fan, Wenyu Song, Thomas G. Kannampallil, Adam B. Wilcox, Linying Zhang

**Author notes:** Corresponding author: Linying Zhang, Institute for Informatics, Data Science and Biostatistics, Washington University School of Medicine in St. Louis, 660 South Euclid Ave, St. Louis, MO, USA, 63110.

## Abstract

**Objective:** This study investigated associations between social determinants of health (SDoH) and time to MOUD initiation across care settings, providing insights for targeted interventions to promote equitable care for OUD patients.

**Material and Methods:** We linked patient-level electronic health records (EHRs) from a regional integrated health system with census-tract–level SDoH from the Population Level Analysis and Community Estimates (PLACES) database. The study cohort included patients newly diagnosed with OUD (including overdose) between 2000 and 2024. We assessed temporal trends in newly diagnosed OUD cases and MOUD prescriptions, mapped the spatial correlation between OUD cases and the Area Deprivation Index, and used multivariable regression models to quantify associations between SDoH and MOUD initiation, adjusting for demographics, insurance type, and comorbidities. Analyses were stratified by care setting (emergency department, inpatient, and outpatient) to examine setting-specific associations.

**Results:** During 2000-2024, 51,521 patients with OUD or opioid overdose, among whom 14,858 (28.8%) received MOUD. OUD diagnoses peaked at 3,787 cases in 2017, then declined by 42.1% to 2,191 cases in 2024. MOUD initiation, especially buprenorphine, steadily increased throughout the study period. Geospatial analyses revealed more OUD cases in high-ADI neighborhoods. In multivariate analyses, older age and Black or African American race were associated with slower MOUD initiation. In the stratified analyses by care setting, significant associations between SDoH and MOUD initiation were primarily observed in the outpatient setting, though effect sizes were modest.

**Conclusions:** Integrating neighborhood-level SDoH with EHRs can uncover care-setting-specific disparities in treatment initiation and identify neighborhoods with unmet treatment needs.

## Introduction

Opioid use disorder (OUD) and opioid overdose represent urgent public health crises in the United States.^1^ From 2019 to 2023, opioid-involved overdose deaths increased from 49,860 to 79,358, accounting for 75.6% of all overdose deaths nationwide in 2023. The burden of opioid-involved overdose death is uneven across racial and ethnic groups: in 2023, the age-adjusted death rate for opioid-involved overdose death was 37.6 per 100,000 among non-Hispanic Black persons, compared with 23.2 per 100,000 among non-Hispanic White persons.^1^

Medications for opioid use disorder (MOUD)—including methadone,^2–7^ buprenorphine,^8–12^ and naltrexone^13–17^—are shown to reduce morbidity and mortality among patients with OUD by both clinical trials and large-scale observational studies.^18,19^ Despite their proven effectiveness, MOUD are highly underutilized in current clinical practice – only about 1 in 5 (22%) patients with past-year OUD received MOUD.^20^ Delays in initiation and lack of treatment can result in missed opportunities to reduce adverse outcomes, including fatal overdoses, HIV infections, and poorer quality of life.^21–23^

Multiple barriers may contribute to this treatment gap.^24^ At the patient level, lack of knowledge, stigma, competing priorities, transportation difficulties, unstable housing, or concern over cost may impede timely initiation.^25,26^ At the provider level, clinicians may lack training or comfort prescribing MOUD, or may perceive regulatory or logistical constraints (e.g., waiver requirements, insufficient linkage to follow-up).^27,28^ At the system level, fragmentation across care settings, limited access to specialty addiction services, poor care coordination, and geographic disparities in provider availability may further delay treatment.^29^ In addition, social determinants of health (SDoH)—the nonmedical conditions in which people live, work, and age—may influence access, uptake, and timing of MOUD initiation.

A growing body of literature has documented that SDoH influence many aspects of health care delivery for OUD.^30–32^ Studies have linked neighborhood disadvantage, poverty, educational attainment, housing instability, and unemployment to worse outcomes across a range of conditions.^33–35^ In the domain of OUD, however, relatively few studies have directly examined how SDoH relate to MOUD initiation, and the existing evidence is mixed.^32^ For example, a recent study among people with HIV and OUD found that each additional favorable SDoH indicator was associated with a 25% increase in the likelihood of MOUD initiation over a six-month follow-up period.^36^ Other work correlating social variables (e.g., income, housing, criminal justice involvement) with OUD treatment outcomes suggests that disadvantage may impede engagement and retention, but these studies often focus on broad outcome measures rather than the timing of initiation.^37,38^

Separately, the type of care setting through which a patient presents may substantially affect the likelihood and speed of MOUD initiation. For example, initiation in the ED is driven by induction protocols and warm handoffs, inpatient settings utilize addiction consult services, and outpatient settings leverage opioid treatment program (OTP) capacity and follow-up access.^39–42^ Medication-specific requirements further interact with these workflows— methadone is dispensed through federally regulated OTPs, buprenorphine can be prescribed in office-based settings, and extended-release naltrexone requires a period of abstinence prior to initiation. These care settings also differ in workflow constraints, risk tolerance, available staff, and reasons for patient presentation, all of which could potentially moderate how SDoH influence initiation patterns.^24,43^ Understanding how SDoH intersects with these care settings is therefore essential for designing targeted interventions, allocating resources, and monitoring equity at the health-system level.

Yet routine health-system measurement of SDoH remains limited. Individual-level social risks are inconsistently captured in structured electronic health records (EHRs), hindering surveillance and equity-focused quality improvement.^44–46^Because a patient’s home address is often recorded in the EHR, linking patient records to neighborhood-level geospatial data provides a scalable, system-wide proxy for social context that supports equity-focused monitoring and quality improvement.^47^ Prior studies have demonstrated the feasibility of such linkages, but few have leveraged them to quantify care-setting-specific disparities in treatment initiation, especially initiation of MOUD, which can lead to more actionable implementation strategies to improve equity within a health system.^48,49^

Given this context, we sought to better understand the associations between SDoH and time to MOUD initiation, and whether these associations differ by care setting. Our results aim to reveal setting-specific disparities in the initiation of MOUD, highlight populations at risk for delayed initiation, and inform targeted interventions to promote equity in MOUD delivery.

## Methods

### Study Population

This study included OUD (or overdose) patients from Washington University School of Medicine or BJC Health System who had ≥1 corresponding diagnosis code in any care setting from 2000-2024. Due to location-history data is only available since mid-2019, we identified “recently and newly diagnosed” patients whose first diagnosis of OUD or opioid overdose was between January 1, 2020, and December 31, 2024 (Figure S1) to investigate associations between neighborhood-level SDoH and (i) OUD and opioid overdose and (ii) time to MOUD initiation. Patients were also required to have at least one year of continuous observation in the EHR prior to first OUD or opioid overdose diagnosis to ensure that the OUD or overdose diagnosis represents a newly diagnosed case and to allow adequate ascertainment of baseline comorbidities. Care setting was defined as the care setting associated with the first OUD or overdose diagnosis (i.e., emergency department (ED), inpatient, or outpatient) and used to stratify the cohort for care-setting–specific analyses. The medical codes used for phenotyping the study population are available in Supplementary Materials (Table S3 and Table S4).

### Outcomes

The outcome was defined as the days from first OUD (or overdose) diagnosis to first MOUD prescription in the patient’s EHR. The follow-up period was 365 days after diagnosis date. Patients who did not receive MOUD during the follow-up period were considered to be censored at their last recorded clinical encounter or the end of the study period, whichever was earlier.

### Data Sources

Data for this study was retrieved from all clinical encounters at Washington University School of Medicine and BJC Health System between 01/01/2000 and 12/31/2024. These hospitals and clinics (17 hospitals and 3 clinics) are primarily located in the greater St. Louis, southern Illinois and mid-Missouri regions, serving the health care needs of urban, suburban, and rural communities. All EHR data were transformed to the Observational Medical Outcomes Partnership (OMOP) Common Data Model (CDM) version 5.3,^50^ except data on location history, which remain in Epic Clarity data model and were only available since mid-2019 onward, when BJC Health System switched to the Epic EHR system. This study was approved by the Institutional Review Board of Washington University in St. Louis (Protocol #202406102) with a waiver of informed consent.

The SDoH factors were extracted from Population Level Analysis and Community Estimates (PLACES) database.^51^ PLACES, developed by the Centers for Disease Control and Prevention (CDC) in collaboration with the Robert Wood Johnson Foundation (RWJF) and the CDC Foundation, provides estimates for over 40 health indicators, including *9* social determinants of health (SDoH) derived from the 5-year American Community Survey (ACS). PLACES provides data at the county, place, census tract, and ZIP Code Tabulation Area levels across the United States.

The ADI information was obtained from the Neighborhood Atlas, developed by University of Wisconsin School of Medicine and Public Health.^52^ The ADI represents a composite measure that incorporates 17 socioeconomic indicators in the domains of poverty, education, housing, and employment at the census block group level. This validated index has been extensively used in health disparity research and provides a complete assessment of neighborhood disadvantages.

### Covariates

We include the following covariates in the study: 1) Demographic characteristics, such as sex (male, female), age group at the time of initial OUD diagnosis (<18, 18-34, 35-49, 50-64, and >64 years), ethnicity (Hispanic or Latino, Not Hispanic or Latino, and Other), and race (White, Black or African American, Other), where the race category “Other” includes American Indian or Alaska Native, Asian, Native Hawaiian or Other Pacific Islander, and Unknown. 2) Insurance type associated with the diagnosis visit, which was classified into Medicare, Medicaid, Private Insurance, or Other Insurance. 3) Commodities within 365 days prior to the diagnosis date. Four categories of comorbidities were included due to their prevalence among OUD and opioid overdose patients: substance use disorder (alcohol use disorder, cannabis use disorder, and tobacco use disorder), chronic pain, mental disorders (depression, bipolar disorder, schizophrenia), and infectious diseases (human immunodeficiency virus, hepatitis C virus, hepatitis B virus).^53^ 4) All available SDoH from the PLACES database and the ADI from the Neighborhood Atlas.

### Geocoding and Geospatial Data Integration

We implemented an end-to-end pipeline that integrates geospatial data with EHRs (Figure 1).^54^ The pipeline begins with geocoding patient addresses from the EHR using an institutional HIPAA-compliant ArcGIS server to convert addresses to geographic coordinates (latitude and longitude)^21^.

**Figure 1.**
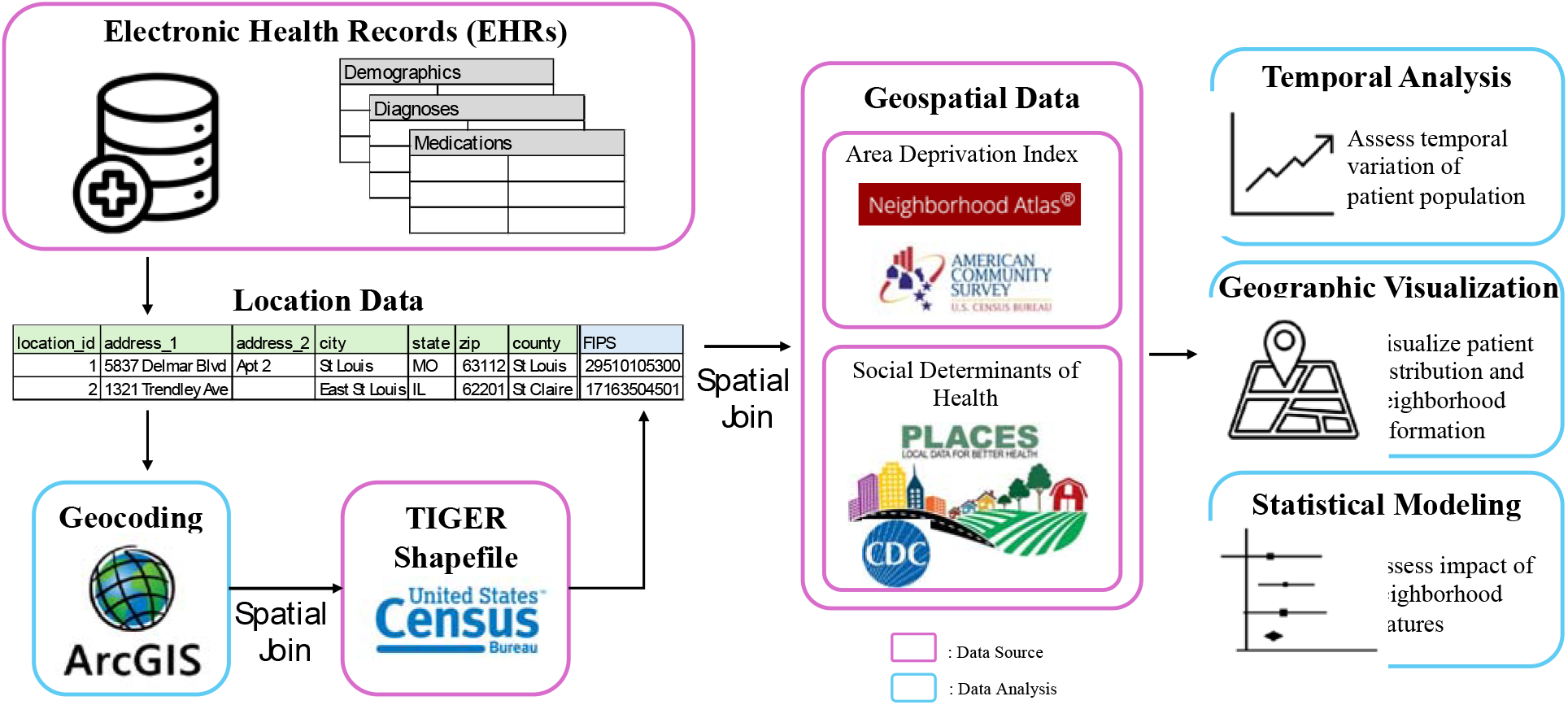
The workflow of integrating census data with electronic health records for health disparity research.

Geocoding quality was assessed using the ArcGIS match score (0–100). The geographic coordinates are then spatially joined to the 2020 US Census Bureau TIGER/Line shapefiles to assign Federal Information Processing Series (FIPS) codes.^55^ Finally, the geocoded EHR data are linked with external geospatial datasets (i.e., PLACES and Neighborhood Atlas) to append census-tract-level ADI and SDoH. The data integration process was performed within the WashU-licensed Azure Databricks cloud environment to best protect patient privacy. The enriched dataset includes ADI and SDoH at the census-tract level, in addition to standard EHR variables (e.g., demographics, diagnoses, medications, and insurance). For this study, we used each patient’s location at the time of the first OUD or opioid overdose diagnosis, so the neighborhood-level ADI and SDoH reflect the social context of the patient at the time of diagnosis.

### Statistical Analysis

We first examined the temporal trends of OUD diagnoses from 2000 to 2024, stratified by sex, race, and age group to assess differential disease burden by demographic subgroups. Second, we created geospatial maps to visualize the geospatial distribution of OUD and opioid overdose and its spatial correlation with ADI.

To examine the associations between neighborhood-level SDoH and time to MOUD initiation within two years following initial OUD or opioid overdose diagnosis, we conducted both unadjusted (i.e. univariate) and adjusted Cox proportional hazards regression analyses with two follow-up periods: 180 days and 365 days. Adjusted analyses included demographics, insurance, and clinical comorbidities in the adjustment set. To assess the association by care setting, we stratified the cohort by care setting (i.e., emergency department, inpatient, and outpatient) at MOUD initiation. Hazards ratio (HR) was reported as a measure of strength of association between the corresponding covariate and MOUD initiation. Statistical analyses were conducted in R.

## Results

From 2000 to 2024, 51,521 patients (mean age 40.64 years; 44.63% female) were newly diagnosed with OUD or opioid overdose, among whom 14,858 (28.8%) patients received at least one prescription of MOUD. For the newly diagnosed subcohort between 2020 and 2024, 10,521 patients were newly diagnosed of OUD or opioid overdose, among whom 3,033 (28.8%) patients received at least one prescription of MOUD. Among those initiating MOUD, 512 (25.35%) initiated in the outpatient setting, 1,340 (32.36%) initiated in the inpatient setting, and 993 (28.06%) in emergency department setting. The Kaplan-Meier curves for time to first MOUD initiation by care setting are shown in Supplementary Materials (Figure S3).

### Temporal Trends of OUD, Opioid Overdose, and MOUD Prescription

As shown in Figure 2, the number of patients diagnosed with OUD or opioid overdose steadily increased from 2000, peaking around 2016-2017 before declining in recent years. Specifically, cases rose sharply after 2010, reaching a peak of 3,787 in 2017, followed by a gradual decline through 2024 to 2,191, representing about a 42.1% decline.

**Figure 2.**
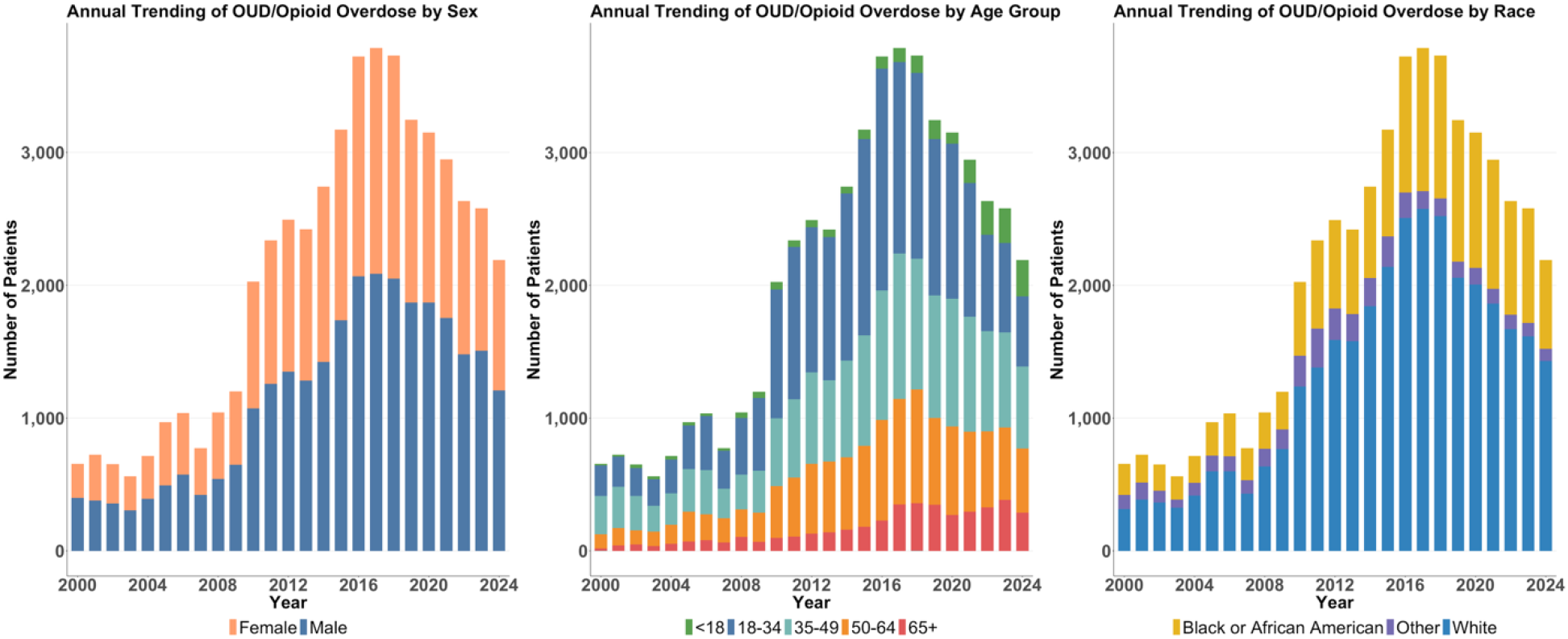
Temporal trends in opioid use disorder (OUD) and opioid overdoes among patients in BJC HealthCare from 2000 to 2024, stratified by sex, age group, race and ethnicity.

Sex-stratified trends indicated that males consistently exhibited higher rates of OUD compared to females, although the gap narrowed slightly over time. Age-stratified trends revealed that individuals aged 18–34 consistently had the highest number of OUD cases throughout the study period, followed by those aged 35–49. All age groups showed similar temporal patterns with peaks around 2017, aligning with the overall trend. Race-stratified trends showed White patients accounted for the majority of OUD cases in the local healthcare system, with a sharp increase after 2010 and a peak around 2017.

The annual trending of MOUD prescriptions revealed the increasing trending of MOUD, especially for buprenorphine, after 2017 and reaching a notable peak in 2024 (Figure 3). The increasing trend of buprenorphine follows the elimination of the X-waiver requirement in late 2022 which indicates the potential policy change making the medication more accessible. In combination with the decrease trends of OUD, the sharp increase could suggest the previous barriers to buprenorphine, instead of clinical need in recent year. The growing trend of methadone could imply the continue utility of OTPs for delivering MOUD. Naltrexone remains consistently underutilized across years.

**Figure 3.**
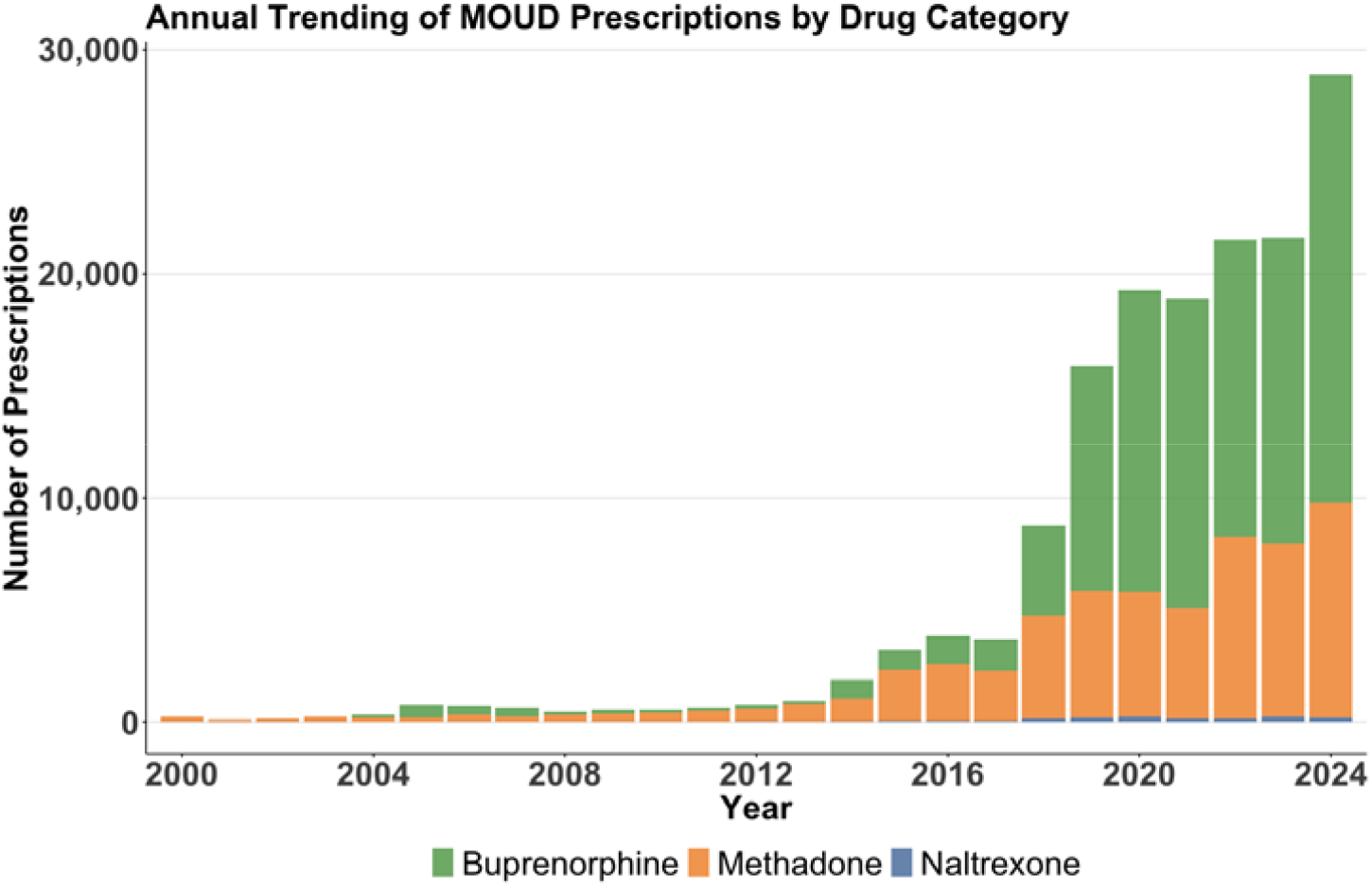
Annual trends in Medication for Opioid Use Disorder prescriptions by drug category among patients in BJC HealthCare from 2000 to 2024. Y-axis represents the number of MOUD prescriptions stratified by medication type (buprenorphine, methadone, and naltrexone).

### Geographical Distribution of OUD and Opioid Overdose

We examined the geographic distribution of OUD and opioid overdose cases and the corresponding untreated population using geographic mapping (Figure 4). The maps illustrate a striking concentration of OUD cases in census tracts with high ADI scores (7–10), which represent the most socioeconomically disadvantaged communities. These high-ADI areas are primarily concentrated within the central, northern and northeastern sectors of the St. Louis metropolitan area. In contrast, low-ADI areas, largely found in the western sectors, show markedly fewer cases. By 2024, high-ADI regions accounted for 57.29% of all OUD cases (n = 4,426) and 63.63% of overdose cases (n = 1,384). In contrast, low-ADI regions represented only 17.51% of OUD cases (n = 1,353) and 15.72% of overdose cases (n = 342) (Figure S2).

**Figure 4.**
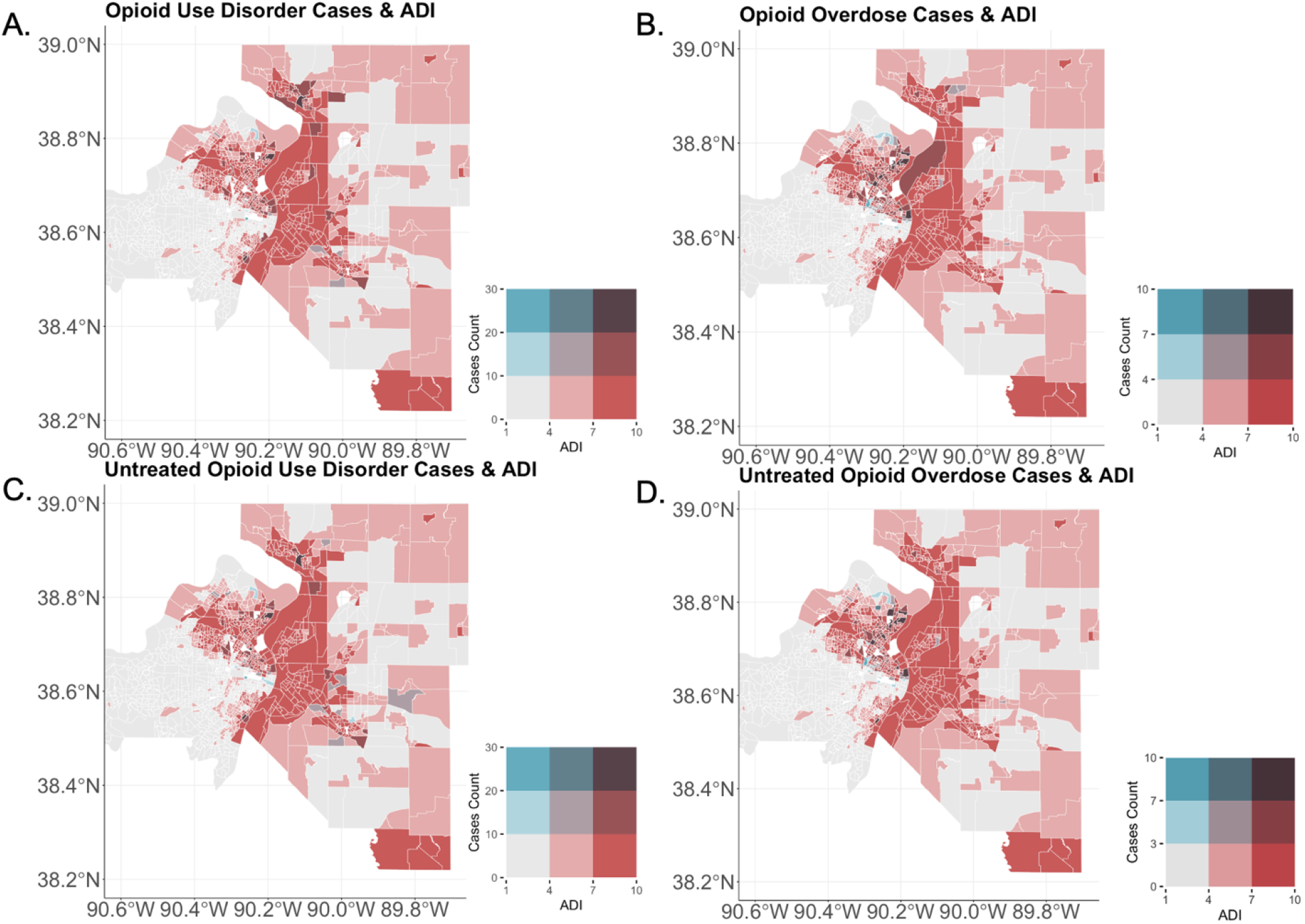
Geographic distribution of opioid use disorder, opioid overdose and untreated patients by census tract in the St. Louis metropolitan area. The bivariate choropleth map displays the intersection of area deprivation index (ADI, red channel) and the number of patients diagnosed with opioid use disorder or opioid overdose (blue channel). Darker shades indicate higher values—greater deprivation (red) and/or more patients (blue). Brown hues reflect areas with both high deprivation and high patient counts.

The geographic overlap between OUD and overdose cases is substantial, especially in the northern sector of the metropolitan area, underscoring a critical area of public health concern. This overlap suggests that specific neighborhoods—many of which fall within the most deprived ADI range—bear a disproportionate burden of opioid-related harms and warrant targeted intervention. Furthermore, a high proportion of untreated patients also resided in these highly deprived areas, rasing the concerns about the potential underutilization of MOUD in these neighborhoods.

### Association between SDoH and MOUD Initiation

Table 1 summarizes patient characteristics by care setting. Inpatient and emergency settings tended to encounter patients from more disadvantaged areas, including those with higher poverty rates and greater housing cost burdens, compared with patients treated in outpatient settings. The univariate associations between SDoH and MOUD initiation is shown in Supplementary Materials (Table S2).

**Table 1.** Characterization of newly diagnosed patients of OUD or opioid overdose from 2020 to 2024, stratified by care setting associated with the diagnosis.

| Characteristic | Care Settings |  |  |
| --- | --- | --- | --- |
|  | Outpatient (N=1,883) | Inpatient (N=3,930) | Emergency (N=3,338) |
| Age Group |  |  |  |
| < 18 | 43 (2.28%) | 126 (3.21%) | 62 (1.86%) |
| 18 - 34 | 457 (24.27%) | 1003 (25.52%) | 1352 (40.50%) |
| 35 - 49 | 507 (26.93%) | 1116 (28.40%) | 1139 (34.12%) |
| 50 - 64 | 477 (25.33%) | 1052 (26.77%) | 596 (17.86%) |
| 65+ | 399 (21.19%) | 633 (16.11%) | 189 (5.66%) |
| Gender |  |  |  |
| Female | 961 (51.04%) | 1729 (43.99%) | 1196 (35.83%) |
| Male | 922 (48.96%) | 2201 (56.01%) | 2142 (64.17%) |
| Race |  |  |  |
| White | 1,402 (74.46%) | 2,456 (62.49%) | 1,784 (53.45%) |
| Black or African American | 460 (24.43%) | 1,381 (35.14%) | 1,485 (44.49%) |
| Other | 21 (1.12%) | 93 (2.37%) | 69 (2.07%) |
| Ethnicity |  |  |  |
| Non-Hispanic or Latino | 1,851 (98.30%) | 3,831 (97.48%) | 3,249 (97.33%) |
| Hispanic or Latino | 14 (0.74%) | 51 (1.30%) | 46 (1.38%) |
| Other | 18 (0.96%) | 48 (1.22%) | 43 (1.29%) |
| Insurance Type |  |  |  |
| Medicare | 160 (8.50%) | 823 (20.94%) | 296 (8.87%) |
| Medicaid | 294 (15.61%) | 1,557 (39.62%) | 1,100 (32.95%) |
| Private Insurance | 90 (4.80%) | 247 (6.28%) | 267 (8.00%) |
| Other Insurance | 1,293 (68.7%) | 1,085 (27.61%) | 1,607 (48.14%) |
| No Insurance | 46 (2.40%) | 218 (5.55%) | 68 (2.04%) |
| Comorbidities |  |  |  |
| Depression | 301 (15.99%) | 519 (13.21%) | 250 (7.49%) |
|  | Outpatient (N=1,883) | Inpatient (N=3,930) | Emergency (N=3,338) |
| Chronic pain | 532 (28.25%) | 897 (22.82%) | 355 (10.64%) |
| Bipolar Disorder | 105 (5.58%) | 222 (5.65%) | 177 (5.30%) |
| Schizophrenia | 50 (2.66%) | 149 (3.79%) | 130 (3.89%) |
| Alcohol Use Disorder | 87 (4.62%) | 261 (6.64%) | 170 (5.09%) |
| Cannabis Use Disorder | 35 (1.86%) | 150 (3.82%) | 109 (3.27%) |
| Tobacco Use Disorder | 4 (0.21%) | 9 (0.23%) | 2 (0.06%) |
| HIV | 16 (0.85%) | 54 (1.37%) | 35 (1.05%) |
| HCV | 74 (3.93%) | 156 (3.97%) | 73 (2.19%) |
| HBV | 3 (0.16%) | 6 (0.15%) | 3 (0.09%) |
| Social Determinants of Health, Mean (SD) |  |  |  |
| Housing crowding | 1.50 (2.08) | 1.65 (2.45) | 1.73 (2.50) |
| House cost burden | 26.77 (11.31) | 29.11 (12.28) | 30.73 (12.45) |
| No internet subscription | 15.80 (9.41) | 17.82 (10.85) | 17.82 (10.85) |
| No high school diploma | 10.40 (6.61) | 11.02 (6.89) | 11.35 (6.95) |
| Persons aged 65+ | 16.46 (5.62) | 16.53 (5.87) | 15.99 (6.03) |
| Persons living below 150% of the poverty level | 24.51 (14.39) | 27.47 (15.61) | 28.82 (15.90) |
| Persons of racial or ethnic minority status | 31.38 (31.55) | 40.99 (34.75) | 46.01 (35.77) |
| Single-parent households | 8.03 (5.97) | 8.98 (6.40) | 9.78 (6.83) |
| Unemployment | 5.66 (4.34) | 6.45 (5.06) | 6.82 (5.25) |
| MOUD Treatment |  |  |  |
| Received any MOUD | 463 (24.59%) | 1,276 (32.47%) | 463 (24.59%) |
| Buprenorphine | 377 (20.02%) | 856 (21.78%) | 377 (20.02%) |
| Methadone | 58 (3.08%) | 349 (8.88%) | 58 (3.08%) |
| Naltrexone | 28 (1.49%) | 71 (1.81%) | 28 (1.49%) |

Table 2 presents the associations between SDoH and MOUD initiation from the multivariate Cox regression with 365-day follow-up period. The associations from 180-day follow-up period are presented in Supplementary Materials (Table S1). Across *all care settings*, time to MOUD initiation varies markedly by age and race, with older adults and Black or African American race were associated with slower MOUD initiation. Patients with Hepatitis C virus (HCV) infection was associated with shorter time to MOUD initiation, while chronic pain was associated with longer time to treatment initiation. Sex, insurance type, and SDoH showed no statistically significant associations.

**Table 2.**
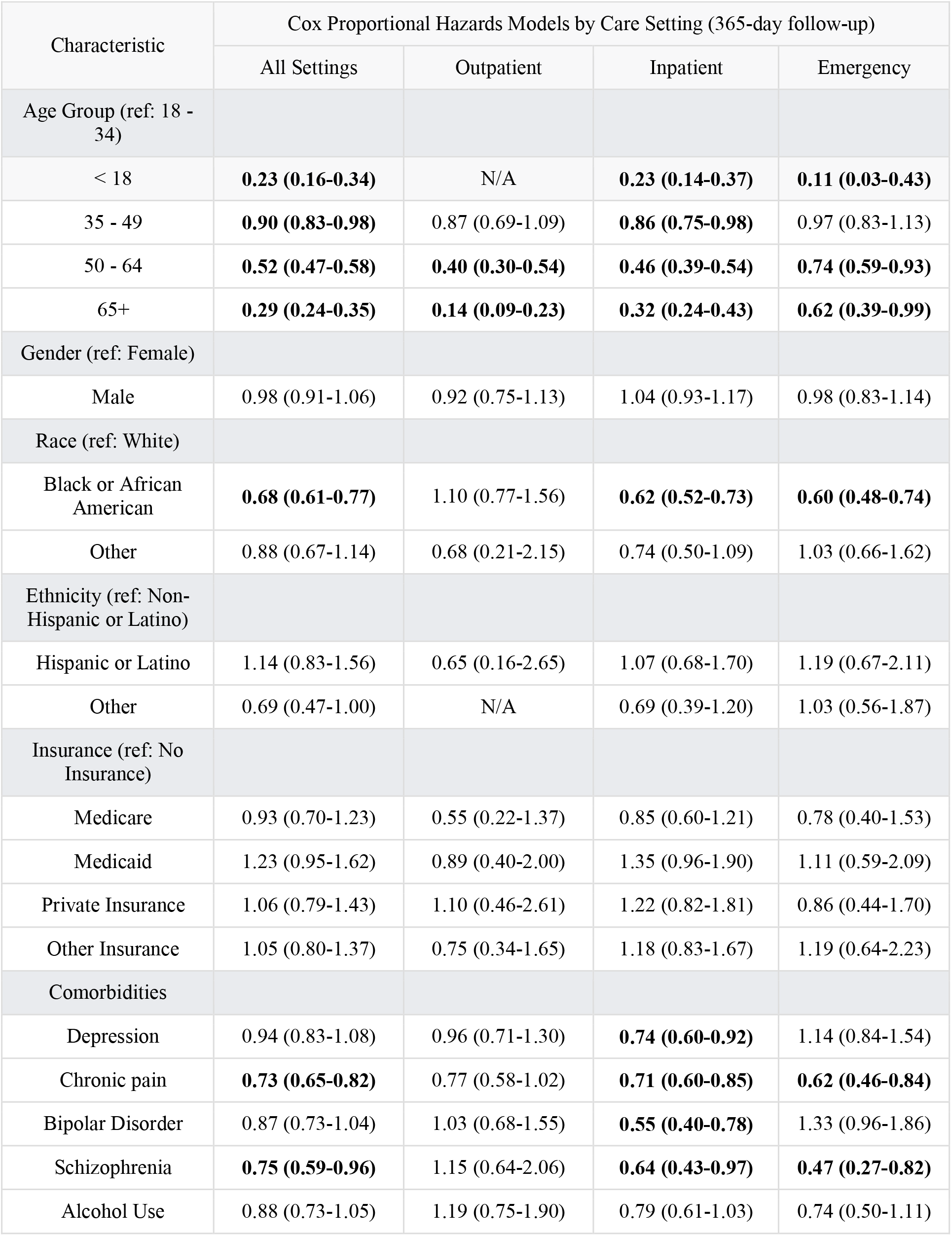
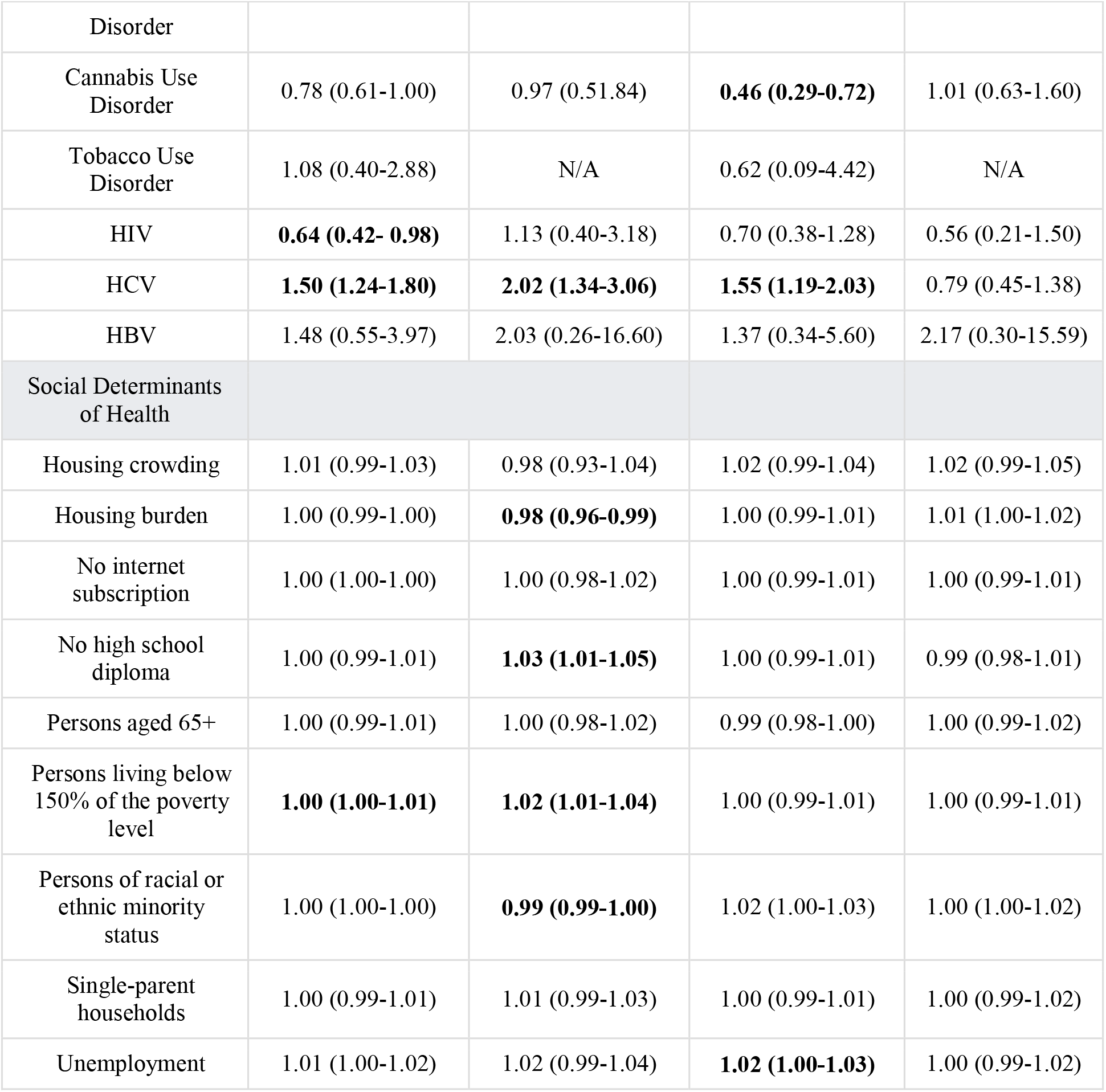
Cox proportional hazards models for time to first MOUD initiation within 365 days of initial OUD or overdose diagnosis, stratified by care setting. Hazard ratios (HRs) with 95% confidence intervals are shown. The HR<1 indicates a longer time from diagnosis to MOUD initiation (i.e., slower initiation). Statistically significant HRs are shown in bold.

| Characteristic | Cox Proportional Hazards Models by Care Setting (365-day follow-up) |  |  |  |
| --- | --- | --- | --- | --- |
|  | All Settings | Outpatient | Inpatient | Emergency |
| Age Group (ref: 18 - 34) |  |  |  |  |
| < 18 | **0.23 (0.16-0.34)** | N/A | **0.23 (0.14-0.37)** | **0.11 (0.03-0.43)** |
| 35 - 49 | **0.90 (0.83-0.98)** | 0.87 (0.69-1.09) | **0.86 (0.75-0.98)** | 0.97 (0.83-1.13) |
| 50 - 64 | **0.52 (0.47-0.58)** | **0.40 (0.30-0.54)** | **0.46 (0.39-0.54)** | **0.74 (0.59-0.93)** |
| 65+ | **0.29 (0.24-0.35)** | **0.14 (0.09-0.23)** | **0.32 (0.24-0.43)** | **0.62 (0.39-0.99)** |
| Gender (ref: Female) |  |  |  |  |
| Male | 0.98 (0.91-1.06) | 0.92 (0.75-1.13) | 1.04 (0.93-1.17) | 0.98 (0.83-1.14) |
| Race (ref: White) |  |  |  |  |
| Black or African American | **0.68 (0.61-0.77)** | 1.10 (0.77-1.56) | **0.62 (0.52-0.73)** | **0.60 (0.48-0.74)** |
| Other | 0.88 (0.67-1.14) | 0.68 (0.21-2.15) | 0.74 (0.50-1.09) | 1.03 (0.66-1.62) |
| Ethnicity (ref: Non-Hispanic or Latino) |  |  |  |  |
| Hispanic or Latino | 1.14 (0.83-1.56) | 0.65 (0.16-2.65) | 1.07 (0.68-1.70) | 1.19 (0.67-2.11) |
| Other | 0.69 (0.47-1.00) | N/A | 0.69 (0.39-1.20) | 1.03 (0.56-1.87) |
| Insurance (ref: No Insurance) |  |  |  |  |
| Medicare | 0.93 (0.70-1.23) | 0.55 (0.22-1.37) | 0.85 (0.60-1.21) | 0.78 (0.40-1.53) |
| Medicaid | 1.23 (0.95-1.62) | 0.89 (0.40-2.00) | 1.35 (0.96-1.90) | 1.11 (0.59-2.09) |
| Private Insurance | 1.06 (0.79-1.43) | 1.10 (0.46-2.61) | 1.22 (0.82-1.81) | 0.86 (0.44-1.70) |
| Other Insurance | 1.05 (0.80-1.37) | 0.75 (0.34-1.65) | 1.18 (0.83-1.67) | 1.19 (0.64-2.23) |
| Comorbidities |  |  |  |  |
| Depression | 0.94 (0.83-1.08) | 0.96 (0.71-1.30) | **0.74 (0.60-0.92)** | 1.14 (0.84-1.54) |
| Chronic pain | **0.73 (0.65-0.82)** | 0.77 (0.58-1.02) | **0.71 (0.60-0.85)** | **0.62 (0.46-0.84)** |
| Bipolar Disorder | 0.87 (0.73-1.04) | 1.03 (0.68-1.55) | **0.55 (0.40-0.78)** | 1.33 (0.96-1.86) |
| Schizophrenia | **0.75 (0.59-0.96)** | 1.15 (0.64-2.06) | **0.64 (0.43-0.97)** | **0.47 (0.27-0.82)** |
| Alcohol Use | 0.88 (0.73-1.05) | 1.19 (0.75-1.90) | 0.79 (0.61-1.03) | 0.74 (0.50-1.11) |
|  | All Settings | Outpatient | Inpatient | Emergency |
| Disorder |  |  |  |  |
| Cannabis Use Disorder | 0.78 (0.61-1.00) | 0.97 (0.51-1.84) | **0.46 (0.29-0.72)** | 1.01 (0.63-1.60) |
| Tobacco Use Disorder | 1.08 (0.40-2.88) | N/A | 0.62 (0.09-4.42) | N/A |
| HIV | **0.64 (0.42- 0.98)** | 1.13 (0.40-3.18) | 0.70 (0.38-1.28) | 0.56 (0.21-1.50) |
| HCV | **1.50 (1.24-1.80)** | **2.02 (1.34-3.06)** | **1.55 (1.19-2.03)** | 0.79 (0.45-1.38) |
| HBV | 1.48 (0.55-3.97) | 2.03 (0.26-16.60) | 1.37 (0.34-5.60) | 2.17 (0.30-15.59) |
| Social Determinants of Health |  |  |  |  |
| Housing crowding | 1.01 (0.99-1.03) | 0.98 (0.93-1.04) | 1.02 (0.99-1.04) | 1.02 (0.99-1.05) |
| Housing burden | 1.00 (0.99-1.00) | **0.98 (0.96-0.99)** | 1.00 (0.99-1.01) | 1.01 (1.00-1.02) |
| No internet subscription | 1.00 (1.00-1.00) | 1.00 (0.98-1.02) | 1.00 (0.99-1.01) | 1.00 (0.99-1.01) |
| No high school diploma | 1.00 (0.99-1.01) | **1.03 (1.01-1.05)** | 1.00 (0.99-1.01) | 0.99 (0.98-1.01) |
| Persons aged 65+ | 1.00 (0.99-1.01) | 1.00 (0.98-1.02) | 0.99 (0.98-1.00) | 1.00 (0.99-1.02) |
| Persons living below 150% of the poverty level | **1.00 (1.00-1.01)** | **1.02 (1.01-1.04)** | 1.00 (0.99-1.01) | 1.00 (0.99-1.01) |
| Persons of racial or ethnic minority status | 1.00 (1.00-1.00) | **0.99 (0.99-1.00)** | 1.02 (1.00-1.03) | 1.00 (1.00-1.02) |
| Single-parent households | 1.00 (0.99-1.01) | 1.01 (0.99-1.03) | 1.00 (0.99-1.01) | 1.00 (0.99-1.02) |
| Unemployment | 1.01 (1.00-1.02) | 1.02 (0.99-1.04) | **1.02 (1.00-1.03)** | 1.00 (0.99-1.02) |

In *outpatient* setting, older adults initiated treatment slower. Compared with people in the 18-34 age group, patients aged 50–64 years (aHR 0.40, 95% CI 0.30-0.54) and 65+ years (aHR 0.14, 95% 0.09-0.23) had longer time to treatment. HCV infection significantly shortened the treatment initiation time (aHR 2.02, 95% CI 1.34-3.06). No race significant difference was observed in outpatient setting. Outpatient initiation showed the most pronounced SDoH influence: higher housing cost burden was associated with slower initiation (aHR 0.98, 95% CI 0.96–0.99); neighborhoods with a greater share of racial/ethnic minority residents also showed slower initiation (aHR 0.99, 95% CI 0.99–1.00). Conversely, greater poverty (aHR 1.02, 95% CI 1.01–1.04) and lower educational attainment (aHR 1.03, 95%CI 1.01–1.05) were associated with faster outpatient initiation.

In *inpatient* setting, all age groups had slower treatment imitation compared with 18-34 age group. Black or African American patients had substantially slower initiation in inpatient settings (aHR 0.62, 95%CI 0.52-0.73). MOUD initiation was most strongly associated with several psychiatric and pain comorbidities. Slower initiation was associated with depression (aHR 0.74, 95% CI 0.60-0.92), chronic pain (aHR 0.71, 95%CI 0.60-0.85), bipolar disorder (aHR 0.55, 95%CI 0.40-0.78), schizophrenia (aHR 0.64, 95% CI 0.43-0.97) and cannabis use disorder (aHR 0.46, 95% CI 0.29–0.72). In contrast, HCV was strengthened after multivariable adjustment and associated with faster initiation in inpatient (aHR 1.55, 95% CI 1.19–2.03). Insurance types were no longer significant once adjusted for the other variables. The only SDoH that is significant associated is unemployment rate (aHR 1.02, 95% CI 1.00-1.03).

In *emergency* setting, 50-64 years older patients (aHR 0.75, 95% CI 0.59-0.95) and Black American patients (aHR 0.60, 95% 0.48–0.75) still had delays in treatment initiation. Only patients with chronic pain (aHR 0.62, 95% CI 0.46-0.84) and schizophrenia (aHR 0.47, 95% CI 0.27-0.82) had slower initiation. SDoH associations in the emergency setting were small and not statistically significant.

## Discussion

Based on a large-scale study, we found that the OUD or opioid overdose cases were more prevalent in socioeconomically disadvantaged neighborhoods, where a higher proportion of patients also remained untreated. Timely MOUD initiation was differentially associated with patient demographics, comorbidities, and SDoH across clinical settings. Neighborhood SDoH were most strongly associated with outpatient initiation, inpatient initiation aligned more closely with clinical comorbidity burden, and ED initiation was not affected by most of the clinical comorbidities and SDoH after adjustment.

### Interpretation in health-system context

Outpatient initiation depends on a sequence of patient- and system-facing steps—scheduling, transportation, childcare, work flexibility, pharmacy access, and follow-up—each of which is sensitive to neighborhood-level disadvantage (e.g., housing cost burden, limited transit, broadband gaps).^56,57^ Even with reduced regulatory barriers for buprenorphine, outpatient initiation often requires reliable phone/internet contact and timely appointments; housing instability and competing social needs can delay or derail these steps. In contrast, inpatient initiation typically unfolds within a highly protocolized, time-bounded episode dominated by clinical acuity and medication reconciliation. Here, psychiatric comorbidity and chronic pain—common reasons for hospitalization—may complicate assessment for withdrawal risk, medication interactions, or sedation, leading to deferment of initiation until post-discharge.^58,59^ ED care, focused on stabilization and rapid disposition, offers a narrow window for initiation and social context may have less opportunity to affect the immediate decision to start, though it likely influences post-ED engagement. The faster initiation observed in patients with HCV is consistent with integrated infectious-disease–addiction pathways and clinical prioritization for high-risk patients.^53^

A note on counterintuitive outpatient signals (e.g., faster initiation in higher-poverty tracts and areas with low educational attainment): these may reflect (i) service-location effects (clinics or outreach programs colocated in high-poverty neighborhoods), (ii) differential triage that channels patients from disadvantaged tracts into low-threshold or walk-in pathways, or (iii) residual facility/clinician confounding not captured in our models.

### Implications for targeted implementation

Findings in this study can be translated into setting-specific design targets. In outpatient setting, health systems could consider expanding low-threshold access (walk-in, evening/weekend hours, telehealth starts), colocating services in high-ADI tracts, and leverage community pharmacies and mobile teams for initiation and early follow-up, and investing in practical enablers—ride support, childcare stipends, and phone/data access.

In both ED and inpatient settings, while initiation during the visit is often not feasible due to time or resource constraints, automate appointment scheduling *before* discharge, include MOUD in discharge bundles with pharmacy fulfillment and transportation vouchers, and deploy social workers and addiction recovery coaches to ensure linkage between care visits.

### Limitations

There are several limitations for this study. First, this is a single center study, albeit with a large patient population across a long study period. Second, neighborhood measures act as proxies for individual social risks and may be inaccurate. Third, location history was available only in recent years, so the findings may not generalize to earlier years. Fourth, MOUD received outside the health system (e.g., methadone dispensed at OTPs) were not captured in the EHR, leading to potentially misclassified initiation timing and setting. Fifth, competing events, such as deaths, incarceration, or migration can lead to censoring that is not completely noninformative. Fifth, interaction between SDoH and other factors are not accounted for in this study. Finally, our findings reflect a single regional health system and may not generalize to other health care systems.

## Conclusion

Linking geospatial context to EHRs provides a practical, scalable approach for care-setting–specific equity surveillance and intervention design. In this regional health system, neighborhood social disadvantage most strongly hindered outpatient initiation, while inpatient timing was driven by clinical complexity and emergency department initiation showed limited sensitivity to SDoH after adjustment. The distinct drivers of care delivery revealed in this study call for care-setting-specific implementations to accelerate timely, equitable MOUD initiation and reduce opioid-related harms.

## Data Availability

Due to ethical restrictions related to patient privacy, the raw data cannot be shared publicly.

## Acknowledgments

This work was supported by internal funding from the Washington University Transdisciplinary Institute in Applied Data Sciences (TRIADS) Seed Grant Program (PJ000030883) and the Washington University Here and Next Research Grant (PJ000030799). The authors would like to acknowledge Shinji Naka and Snehil Gupta from the WashU Informatics Core Services (ICS) for their guidance on querying EHR data.

