## Supplementary Materials for "Differential Associations Between Social Determinants of Health and the Initiation of Medications for Opioid Use Disorder Across Care Settings"

### **Figure S1.** Flow diagram of the participants selection and classification criteria for geospatial cohort.

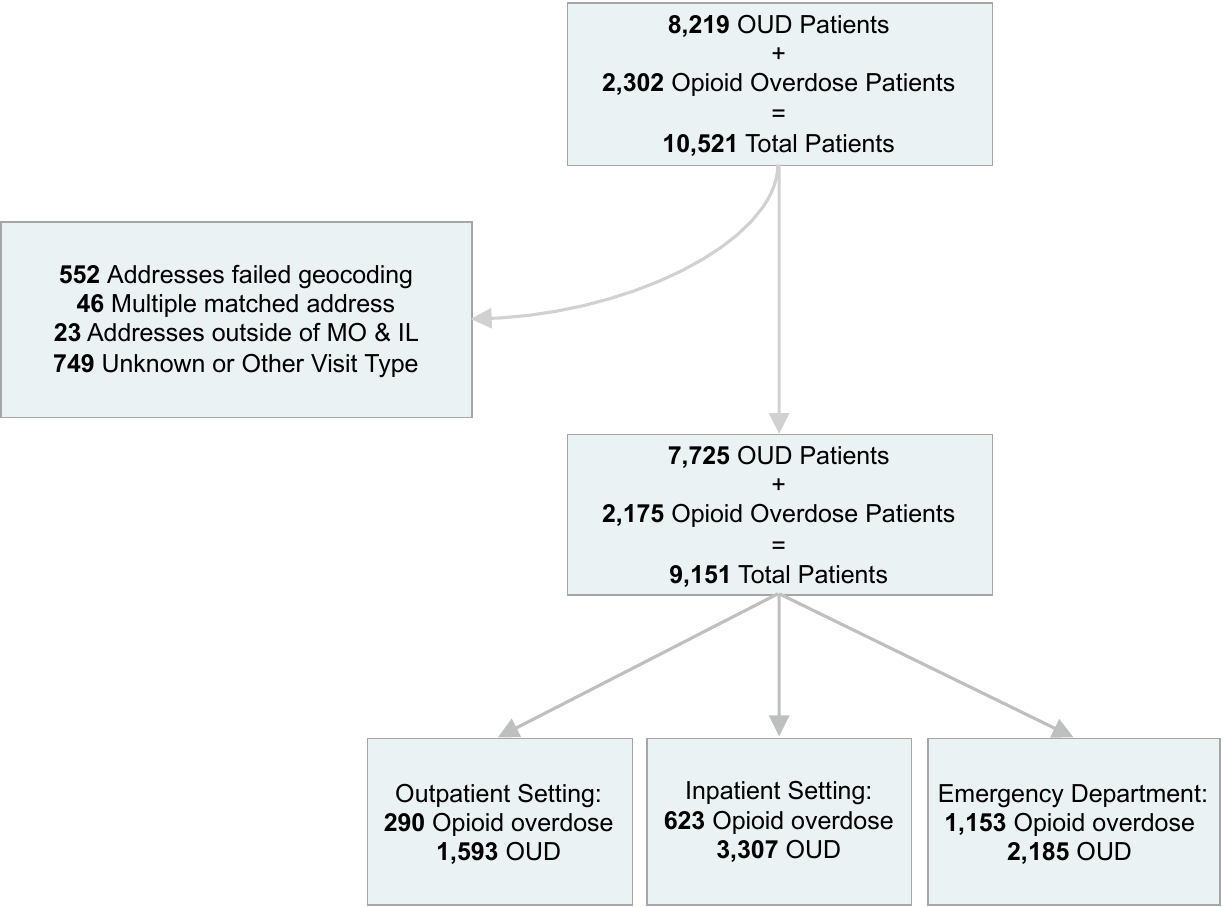
We matched each patient's first OUD or opioid overdose diagnosis date to the nearest location record, with a mean temporal difference of 87.93 days (SD: 160.33).

*OUD = Opioid Use Disorder

### **Figure S2.** Cumulative incidence of OUD and opioid overdose cases from 2020 to 2024, stratified by state-level Area Deprivation Index (ADI) tertiles.

ADI is classified into three categories: 1st tertile (1-3) represents the least deprived areas, 2nd tertile (4-6) represents moderately deprived areas, and 3rd tertile (7-10) represents the most deprived areas.

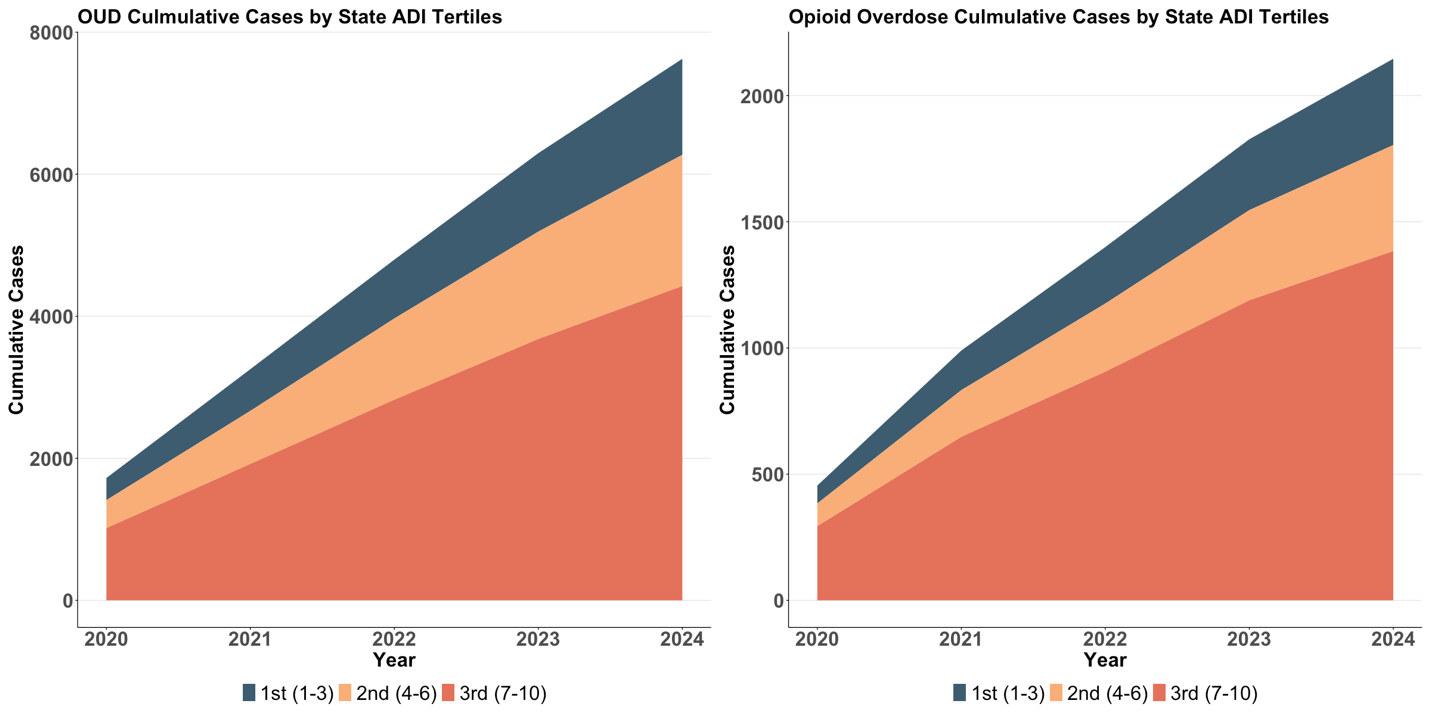

*ADI = Area Deprivation Index

*OUD = Opioid Use Disorder

### **Figure S3.** Kaplan–Meier curves for time to first MOUD initiation by care setting among patients newly diagnosed with opioid use disorder between 2020 and 2024.

Curves represent the probability of not receiving MOUD treatment over 60 days following OUD or opioid overdose diagnosis with inpatient settings showed the fastest treatment initiation (log-rank test p < 0.0001).

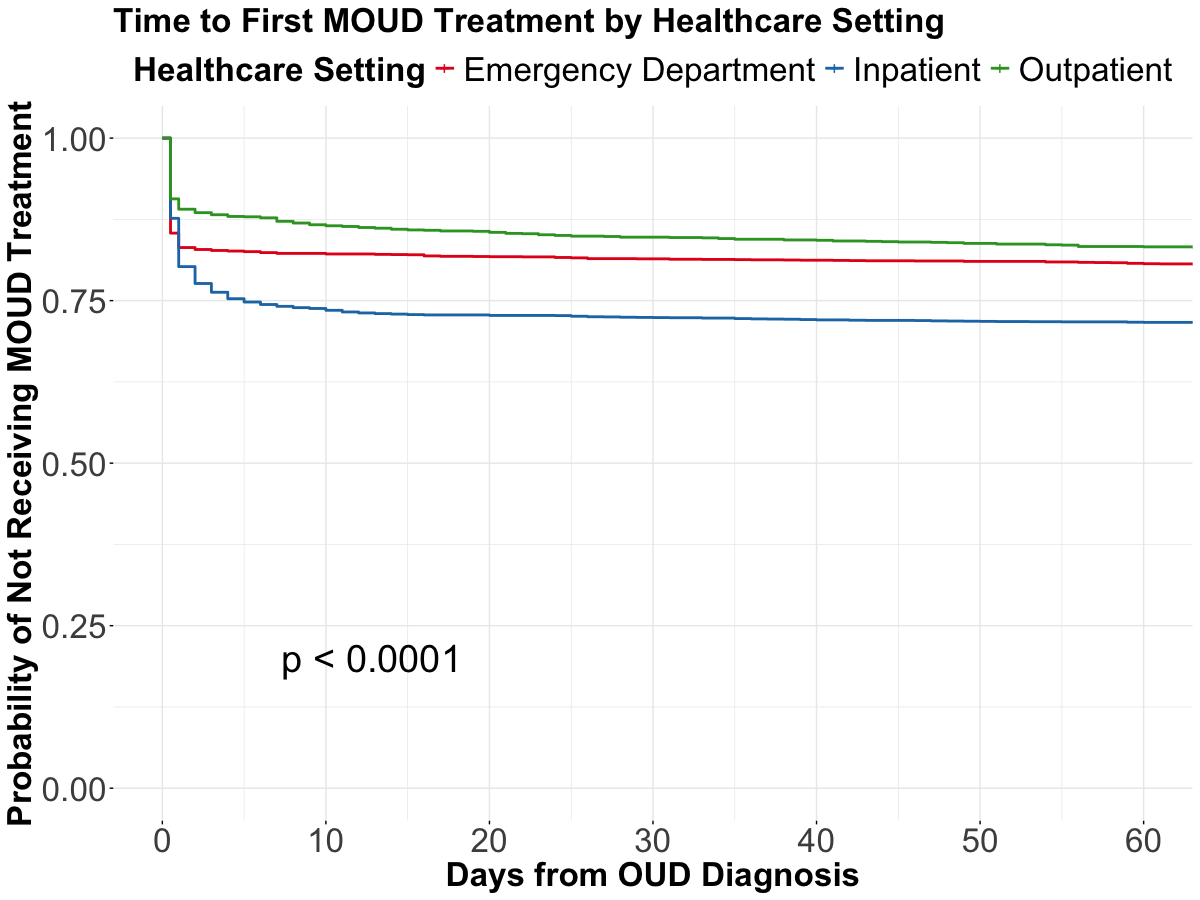

*MOUD = Medication to Opioid Use Disorder

*OUD = Opioid Use Disorder

### **Table S1.** Cox proportional hazards models for time to first MOUD initiation stratified by care setting among patients with opioid use disorder or opioid overdose (180-day follow-up).

Hazard ratios (HRs) with 95% confidence intervals are shown. The HR<1 indicates that the characteristic increases the time from OUD diagnosis to MOUD initiation (i.e., longer time to initiation).

| Characteristic | Cox Proportional Hazards Models by Care Setting (180-day follow-up) | | |
| --- | --- | --- | --- |
|  | Outpatient | Inpatient | Emergency |
| Age Group (ref: 18 - 34) |  |  |  |
| < 18 | N/A | **0.22 (0.13-0.36)** | **0.06 (0.01-0.41)** |
| 35 - 49 | 0.90 (0.71-1.14) | **0.85 (0.74-0.97)** | 0.95 (0.81-1.12) |
| 50 - 64 | **0.41 (0.30-0.55)** | **0.46 (0.39-0.55)** | **0.75 (0.59-0.95)** |
| 65+ | **0.16 (0.10-0.26)** | **0.32 (0.24-0.43)** | 0.65 (0.40-1.05) |
| Gender (ref: Female) |  |  |  |
| Male | 0.92 (0.74-1.14) | 1.05 (0.93-1.18) | 0.98 (0.84-1.15) |
| Race (ref: White) |  |  |  |
| Black or African American | 1.04 (0.72-1.50) | **0.61 (0.51-0.72)** | **0.60 (0.48-0.75)** |
| Other | 0.75 (0.23-2.37) | 0.73 (0.49-1.08) | 1.00 (0.62-1.60) |
| Ethnicity (ref: Non-Hispanic or Latino) |  |  |  |
| Hispanic or Latino | 0.69 (0.17-2.83) | 1.10 (0.70-1.75) | 1.28 (0.72-2.28) |
| Other | N/A | 0.71 (0.41-1.24) | 1.01 (0.54-1.91) |
| Insurance (ref: No Insurance) |  |  |  |
| Medicare | 0.61 (0.23-1.61) | 0.83 (0.57-1.20) | 0.72 (0.37-1.42) |
| Medicaid | 0.94 (0.40-2.24) | 1.41 (0.99-2.01) | 1.03 (0.55-1.94) |
| Private Insurance | 1.30 (0.52-3.25) | 1.28 (0.85-1.92) | 0.77 (0.39-1.53) |
| Other Insurance | 0.83 (0.36-1.93) | 1.24 (0.86-1.78) | 1.08 (0.58-2.03) |
| Comorbidities |  |  |  |
| Depression | 0.97 (0.71-1.33) | **0.69 (0.55-0.87)** | 1.13 (0.82-1.56) |
| Chronic pain | **0.74 (0.55-1.00)** | **0.71 (0.59-0.85)** | **0.59 (0.43-0.82)** |
| Bipolar Disorder | 1.13 (0.75-1.71) | **0.53 (0.37-0.76)** | 1.26 (0.88-1.80) |
| Schizophrenia | 1.05 (0.56-1.97) | **0.64 (0.42-0.98)** | **0.43 (0.23-0.78)** |
| Alcohol Use Disorder | 1.17 (0.72-1.91) | **0.73 (0.56-0.97)** | 0.74 (0.48-1.13) |
| Cannabis Use Disorder | 0.93 (0.47-1.83) | **0.45 (0.28-0.72)** | 1.02 (0.63-1.67) |
| Tobacco Use Disorder | N/A | 0.67 (0.09-4.80) | N/A |
| HIV | 1.27 (0.45-3.55) | 0.74 (0.41-1.36) | 0.47 (0.15-1.48) |
| HCV | **2.05 (1.34-3.13)** | **1.59 (1.21-2.08)** | 0.73 (0.40-1.33) |
| HBV | 2.19 (0.27-17.52) | 1.46 (0.36-5.99) | N/A |
| Social Determinants of Health |  |  |  |
| Housing crowding | 0.99 (0.94-1.04) | 1.02 (0.99-1.04) | 1.02 (0.98-1.05) |
| House cost burden | **0.98 (0.96-0.99)** | 1.00 (0.99-1.01) | 1.01 (1.00-1.02) |
| No internet subscription | 1.00 (0.99-1.02) | 1.00 (0.99-1.01) | 1.00 (0.99-1.01) |
| No high school diploma | **1.03 (1.01-1.05)** | 1.00 (0.99-1.01) | 0.99 (0.98-1.01) |
| Persons aged 65+ | 1.00 (0.98-1.02) | 0.99 (0.98-1.00) | 1.00 (0.99-1.02) |
| Persons living below 150% of the poverty level | **1.02 (1.00-1.04)** | 1.00 (0.99-1.01) | 1.00 (0.99-1.01) |
| Persons of racial or ethnic minority status | **0.99 (0.99-1.00)** | 1.00 (1.00-1.00) | 1.00 (1.00-1.00) |
| Single-parent households | 1.01 (0.99-1.03) | 1.00 (0.99-1.01) | 1.00 (0.99-1.02) |
| Unemployment | 1.02 (0.99-1.04) | **1.02 (1.00-1.03)** | 1.00 (0.98-1.02) |

*HIV = Human immunodeficiency virus

*HCV = Hepatitis C virus

*HBV = Hepatitis B virus

### **Table S2.** Univariate Cox proportional hazards models for time to first MOUD initiation stratified by care setting among patients with opioid use disorder or opioid overdose (365-day follow-up).

Hazard ratios (HRs) with 95% confidence intervals are shown. The HR<1 indicates that the characteristic increases the time from OUD diagnosis to MOUD initiation (i.e., longer time to initiation).

| Characteristic | Cox Proportional Hazards Models by Care Setting (180-day follow-up) | | |
| --- | --- | --- | --- |
|  | Outpatient | Inpatient | Emergency |
| Age Group (ref: 18 - 34) |  |  |  |
| < 18 | NA | **0.23 (0.14-0.39)** | **0.06 (0.01-0.40)** |
| 35 - 49 | 0.86 (0.68-1.08) | **0.84 (0.73-0.96)** | 0.89 (0.76-1.05) |
| 50 - 64 | **0.38 (0.28-0.51)** | **0.40 (0.34-0.47)** | **0.61 (0.49-0.77)** |
| 65+ | **0.13 (0.08-0.21)** | **0.22 (0.17-0.28)** | **0.45 (0.29-0.68)** |
| Gender (ref: Female) |  |  |  |
| Male | 0.84 (0.68-1.04) | 1.10 (0.98-1.23) | 0.95 (0.82-1.11) |
| Race (ref: White) |  |  |  |
| Black or African American | 0.90 (0.70-1.15) | **0.70 (0.62-0.80)** | **0.63 (0.54-0.73)** |
| Other | 0.71 (0.23-2.21) | 0.94 (0.64-1.36) | 1.11 (0.70-1.76) |
| Ethnicity (ref: Non-Hispanic or Latino) |  |  |  |
| Hispanic or Latino | 0.716 (0.178-2.874) | 1.40 (0.90-2.17) | 1.24 (0.70-2.20) |
| Other | NA | 0.95 (0.55-1.64) | 1.12 (0.60-2.09) |
| Insurance (ref: No Insurance) |  |  |  |
| Medicare | 0.69 (0.27-1.78) | 0.82 (0.56-1.18) | 0.80 (0.40-1.54) |
| Medicaid | 2.09 (0.91-4.80) | **2.38 (1.71-3.32)** | 1.40 (0.74-2.49) |
| Private Insurance | **2.54 (1.05-6.15)** | **2.22 (1.50-3.26)** | 1.09 (0.56-2.10) |
| Other Insurance | 1.45 (0.64-3.23) | **2.20 (1.57-3.08)** | 1.59 (0.87-2.89) |
| Comorbidities |  |  |  |
| Depression | 0.82 (0.61-1.11) | **0.48 (0.39-0.60)** | 0.86 (0.65-1.16) |
| Chronic pain | **0.51 (0.39-0.67)** | **0.45 (0.38-0.53)** | **0.52 (0.38-0.70)** |
| Bipolar Disorder | 1.51 (1.03-2.22) | **0.44 (0.31-0.63)** | 0.96 (0.69-1.33) |
| Schizophrenia | 1.31 (0.74-2.33) | **0.50 (0.33-0.74)** | **0.39 (0.22-0.70)** |
| Alcohol Use Disorder | 1.13 (0.71-1.80) | **0.62 (0.47-0.82)** | **0.61 (0.40-0.91)** |
| Cannabis Use Disorder | 1.53 (0.82-2.87) | **0.36 (0.22-0.57)** | 0.73 (0.46-1.16) |
| Tobacco Use Disorder | NA | 0.34 (0.05-2.40) | NA |
| HIV* | 1.36 (0.51-3.63) | 0.65 (0.36-1.18) | 0.37 (0.12-1.15) |
| HCV^*^ | 2.27 (1.54-3.34) | 1.33 (1.02-1.73) | 0.67 (0.37-1.22) |
| HBV^*^ | 2.11 (0.30-15.00) | 1.12 (0.28-4.49) | NA |
| Social Determinants of Health |  |  |  |
| Housing crowding | **1.05 (1.01-1.10)** | **1.03 (1.01-1.05)** | 1.01 (0.98-1.04) |
| House cost burden | 1.00 (0.99-1.01) | 1.00 (0.99-1.00) | 1.00 (0.99-1.00) |
| No internet subscription | **1.02 (1.01-1.03)** | 1.00 (0.99-1.00) | 1.00 (0.99-1.01) |
| No high school diploma | **1.04 (1.03-1.06)** | 1.00 (0.99-1.01) | 1.00 (0.99-1.01) |
| Persons aged 65+ | 0.99 (0.98-1.01) | **0.99 (0.98-1.00)** | 1.00 (0.99-1.01) |
| Persons living below 150% of the poverty level | **1.02 (1.01-1.02)** | 1.00 (1.00-1.00) | 1.00 (1.00-1.00) |
| Persons of racial or ethnic minority status | 1.00 (0.99-1.00) | **0.998 (0.996-0.999)^*^** | **0.996 (0.994-0.998)^*^** |
| Single-parent households | 1.01 (0.99-1.03) | 1.00 (1.00-1.00) | 1.00 (0.98-1.01) |
| Unemployment | **1.03 (1.00-1.05)** | 1.00 (1.00-1.02) | 0.99 (0.98-1.01) |

*HIV = Human immunodeficiency virus

*HCV = Hepatitis C virus

*HBV = Hepatitis B virus

*Some hazard ratios reported to 3 decimal places to show precision for small effects

### **Table S3:** List of conditions used for cohort construction and included in the multivariate Cox regression model.

Conditions were grouped into the following categories: opioid use disorder, opioid overdose, chronic pain, depression, bipolar disorder, schizophrenia, cannabis use disorder, alcohol use disorder, tobacco use disorder, human immunodeficiency virus (HIV), hepatitis B virus (HBV), and hepatitis C virus (HCV). For each category, descendant concept IDs and names are listed.

| **Concept Id** | **Concept Name** | | **Vocabulary** |
| --- | --- | --- | --- |
| **Opioid Use Disorder** | | | |
| 45766638 | Buprenorphine dependence | | SNOMED |
| 45766639 | Buprenorphine withdrawal | | SNOMED |
| 37165540 | Combined opioid with non-opioid substance dependence | | SNOMED |
| 37165545 | Combined opioid with non-opioid substance dependence, continuous | | SNOMED |
| 37165542 | Combined opioid with non-opioid substance dependence, episodic | | SNOMED |
| 37165546 | Combined opioid with non-opioid substance dependence in remission | | SNOMED |
| 440693 | Continuous opioid dependence | | SNOMED |
| 440379 | Episodic opioid dependence | | SNOMED |
| 4138193 | Fentanyl dependence | | SNOMED |
| 1075432 | Harmful pattern of use of heroin | | SNOMED |
| 1075433 | Harmful pattern of use of intravenous heroin | | SNOMED |
| 4333676 | Heroin dependence | | SNOMED |
| 44782731 | Intravenous nondependent opioid abuse | | SNOMED |
| 4332883 | Methadone dependence | | SNOMED |
| 4338027 | Morphine dependence | | SNOMED |
| 4099935 | Nondependent opioid abuse | | SNOMED |
| 434016 | Nondependent opioid abuse, continuous | | SNOMED |
| 435798 | Nondependent opioid abuse, episodic | | SNOMED |
| 436088 | Nondependent opioid abuse in remission | | SNOMED |
| 438130 | Opioid abuse | | SNOMED |
| 37398751 | Opioid analgesic dependence | | SNOMED |
| 438120 | Opioid dependence | | SNOMED |
| 432301 | Opioid dependence in remission | | SNOMED |
| 42872387 | Opioid dependence, on agonist therapy | | SNOMED |
| 4336384 | Opioid withdrawal | | SNOMED |
| 4332990 | Opium dependence | | SNOMED |
| **Opioid Overdose** | | | |
| 4170820 | Accidental alfentanil overdose | | SNOMED |
| 4156152 | Accidental alfentanil poisoning | | SNOMED |
| 4173535 | Accidental buprenorphine overdose | | SNOMED |
| 4159510 | Accidental buprenorphine poisoning | | SNOMED |
| 4172235 | Accidental dextromoramide overdose | | SNOMED |
| 4156760 | Accidental dextromoramide poisoning | | SNOMED |
| 4172237 | Accidental dextropropoxyphene overdose | | SNOMED |
| 4157359 | Accidental dextropropoxyphene poisoning | | SNOMED |
| 4166625 | Accidental dipipanone overdose | | SNOMED |
| 4157360 | Accidental dipipanone poisoning | | SNOMED |
| 4172243 | Accidental fentanyl overdose | | SNOMED |
| 4156768 | Accidental fentanyl poisoning | | SNOMED |
| 4172240 | Accidental heroin overdose | | SNOMED |
| 4166641 | Accidental levorphanol overdose | | SNOMED |
| 4159523 | Accidental levorphanol poisoning | | SNOMED |
| 4166638 | Accidental meperidine overdose | | SNOMED |
| 4175083 | Accidental meptazinol overdose | | SNOMED |
| 4156156 | Accidental meptazinol poisoning | | SNOMED |
| 4166627 | Accidental methadone overdose | | SNOMED |
| 4173540 | Accidental morphine overdose | | SNOMED |
| 4166632 | Accidental nalbuphine overdose | | SNOMED |
| 4156150 | Accidental nalbuphine poisoning | | SNOMED |
| 4173537 | Accidental overdose by codeine | | SNOMED |
| 4170688 | Accidental overdose by dihydrocodeine | | SNOMED |
| 4178410 | Accidental overdose of opiate | | SNOMED |
| 4166624 | Accidental pentazocine overdose | | SNOMED |
| 4173530 | Accidental phenazocine overdose | | SNOMED |
| 4157358 | Accidental phenazocine poisoning | | SNOMED |
| 4172245 | Accidental phenoperidine overdose | | SNOMED |
| 4157369 | Accidental phenoperidine poisoning | | SNOMED |
| 4313093 | Accidental poisoning by codeine | | SNOMED |
| 4157365 | Accidental poisoning by dihydrocodeine | | SNOMED |
| 440307 | Accidental poisoning by heroin | | SNOMED |
| 438037 | Accidental poisoning by methadone | | SNOMED |
| 4313094 | Accidental poisoning by morphine | | SNOMED |
| 4335394 | Accidental poisoning by opiate agonist | | SNOMED |
| 4311219 | Accidental poisoning by opium | | SNOMED |
| 4313106 | Accidental poisoning by pentazocine | | SNOMED |
| 4311218 | Accidental poisoning by pethidine | | SNOMED |
| 1245119 | Acute opioid intoxication | | SNOMED |
| 4170819 | Alfentanil overdose | | SNOMED |
| 4159517 | Alfentanil poisoning | | SNOMED |
| 4170686 | Buprenorphine overdose | | SNOMED |
| 4157361 | Buprenorphine poisoning | | SNOMED |
| 4173531 | Dextromoramide overdose | | SNOMED |
| 4159505 | Dextromoramide poisoning | | SNOMED |
| 4170680 | Dextropropoxyphene overdose | | SNOMED |
| 4159507 | Dextropropoxyphene poisoning | | SNOMED |
| 4173533 | Dipipanone overdose | | SNOMED |
| 4156762 | Dipipanone poisoning | | SNOMED |
| 4170821 | Fentanyl overdose | | SNOMED |
| 4157368 | Fentanyl poisoning | | SNOMED |
| 4170687 | Heroin overdose | | SNOMED |
| 4166636 | Intentional alfentanil overdose | | SNOMED |
| 4156153 | Intentional alfentanil poisoning | | SNOMED |
| 4166628 | Intentional buprenorphine overdose | | SNOMED |
| 4157362 | Intentional buprenorphine poisoning | | SNOMED |
| 4172236 | Intentional dextromoramide overdose | | SNOMED |
| 4159506 | Intentional dextromoramide poisoning | | SNOMED |
| 4170681 | Intentional dextropropoxyphene overdose | | SNOMED |
| 4159508 | Intentional dextropropoxyphene poisoning | | SNOMED |
| 4054782 | Intentional diamorphine overdose | | SNOMED |
| 4055126 | Intentional dihydrocodeine overdose | | SNOMED |
| 4173534 | Intentional dipipanone overdose | | SNOMED |
| 4159509 | Intentional dipipanone poisoning | | SNOMED |
| 4173541 | Intentional fentanyl overdose | | SNOMED |
| 4159519 | Intentional fentanyl poisoning | | SNOMED |
| 4159511 | Intentional heroin poisoning | | SNOMED |
| 4170826 | Intentional levorphanol overdose | | SNOMED |
| 4157372 | Intentional levorphanol poisoning | | SNOMED |
| 4170823 | Intentional meperidine overdose | | SNOMED |
| 4159521 | Intentional meperidine poisoning | | SNOMED |
| 4172246 | Intentional meptazinol overdose | | SNOMED |
| 4156157 | Intentional meptazinol poisoning | | SNOMED |
| 4055127 | Intentional methadone overdose | | SNOMED |
| 4156147 | Intentional methadone poisoning | | SNOMED |
| 4166634 | Intentional morphine overdose | | SNOMED |
| 4156767 | Intentional morphine poisoning | | SNOMED |
| 4172242 | Intentional nalbuphine overdose | | SNOMED |
| 4156151 | Intentional nalbuphine poisoning | | SNOMED |
| 606804 | Intentional opioid receptor agonist poisoning | | SNOMED |
| 4166630 | Intentional overdose by codeine | | SNOMED |
| 603123 | Intentional overdose of opioid receptor agonist | | SNOMED |
| 4170677 | Intentional pentazocine overdose | | SNOMED |
| 4156757 | Intentional pentazocine poisoning | | SNOMED |
| 4170678 | Intentional phenazocine overdose | | SNOMED |
| 4156758 | Intentional phenazocine poisoning | | SNOMED |
| 4170824 | Intentional phenoperidine overdose | | SNOMED |
| 4156155 | Intentional phenoperidine poisoning | | SNOMED |
| 4157364 | Intentional poisoning by codeine | | SNOMED |
| 4156149 | Intentional poisoning by dihydrocodeine | | SNOMED |
| 4175084 | Levorphanol overdose | | SNOMED |
| 4156158 | Levorphanol poisoning | | SNOMED |
| 4170818 | Meperidine analog overdose | | SNOMED |
| 4159516 | Meperidine analog poisoning | | SNOMED |
| 4170822 | Meperidine overdose | | SNOMED |
| 4166640 | Meptazinol overdose | | SNOMED |
| 4157371 | Meptazinol poisoning | | SNOMED |
| 4172123 | Methadone analog overdose | | SNOMED |
| 4156145 | Methadone analog poisoning | | SNOMED |
| 4170683 | Methadone overdose | | SNOMED |
| 4170685 | Morphinan opioid overdose | | SNOMED |
| 4156764 | Morphinan opioid poisoning | | SNOMED |
| 4166633 | Morphine overdose | | SNOMED |
| 4173539 | Nalbuphine overdose | | SNOMED |
| 4157367 | Nalbuphine poisoning | | SNOMED |
| 4299094 | Opioid intoxication | | SNOMED |
| 4199769 | Opioid intoxication delirium | | SNOMED |
| 4166629 | Overdose of codeine | | SNOMED |
| 4172241 | Overdose of dihydrocodeine | | SNOMED |
| 4053782 | Overdose of opiate | | SNOMED |
| 4172122 | Pentazocine overdose | | SNOMED |
| 4077153 | Phenanthrene derivative poisoning | | SNOMED |
| 4173529 | Phenazocine overdose | | SNOMED |
| 4157357 | Phenazocine poisoning | | SNOMED |
| 4166639 | Phenoperidine overdose | | SNOMED |
| 4156154 | Phenoperidine poisoning | | SNOMED |
| 4194922 | Poisoning by codeine | | SNOMED |
| 4083714 | Poisoning by dihydrocodeine | | SNOMED |
| 433919 | Poisoning by heroin | | SNOMED |
| 4109137 | Poisoning by meperidine | | SNOMED |
| 440919 | Poisoning by methadone | | SNOMED |
| 4166384 | Poisoning by morphine | | SNOMED |
| 37167514 | Poisoning by opium | | SNOMED |
| 4030744 | Poisoning by pentazocine | | SNOMED |
| 606805 | Poisoning caused by opioid receptor agonist | | SNOMED |
| **Chronic Pain** | | | |
| 4044411 | Abdominal cutaneous nerve entrapment syndrome | SNOMED | |
| 40480688 | Acute exacerbation of chronic abdominal pain | SNOMED | |
| 4030546 | Alteration in comfort: chronic pain | SNOMED | |
| 4010966 | Arm claudication | SNOMED | |
| 761828 | Atherosclerosis of bypass graft of bilateral lower limbs with intermittent claudication | SNOMED | |
| 761827 | Atherosclerosis of bypass graft of left lower limb with intermittent claudication | SNOMED | |
| 761826 | Atherosclerosis of bypass graft of right lower limb with intermittent claudication | SNOMED | |
| 35611580 | Bilateral chronic pain following total hip arthroplasty | SNOMED | |
| 36687087 | Bilateral chronic pain of feet | SNOMED | |
| 35615072 | Bilateral chronic pain of upper limbs | SNOMED | |
| 609009 | Bilateral intermittent claudication of lower limbs due to atherosclerosis of nonbiological bypass graft | SNOMED | |
| 37108778 | Bilateral total knee chronic pain following arthroplasty | SNOMED | |
| 193523 | Broad ligament laceration syndrome | SNOMED | |
| 37312248 | Central sensitization | SNOMED | |
| 4008102 | Chronic abdominal pain | SNOMED | |
| 45763561 | Chronic ankle pain | SNOMED | |
| 42537212 | Chronic arthralgia of temporomandibular joint | SNOMED | |
| 4046660 | Chronic back pain | SNOMED | |
| 43530938 | Chronic back pain greater than three months duration | SNOMED | |
| 42538797 | Chronic central neuropathic pain | SNOMED | |
| 37155910 | Chronic central neuropathic pain due to brain injury | SNOMED | |
| 37170539 | Chronic central neuropathic pain due to multiple sclerosis | SNOMED | |
| 37171581 | Chronic central neuropathic pain due to spinal cord injury | SNOMED | |
| 4079008 | Chronic central post-stroke pain | SNOMED | |
| 762941 | Chronic chest pain | SNOMED | |
| 378145 | Chronic cluster headache | SNOMED | |
| 4046226 | Chronic cluster headache evolved from episodic cluster headache | SNOMED | |
| 4047908 | Chronic cluster headache unremitting from onset | SNOMED | |
| 36684905 | Chronic daily headache | SNOMED | |
| 37155935 | Chronic dental pain | SNOMED | |
| 42534971 | Chronic female pelvic pain syndrome | SNOMED | |
| 374639 | Chronic headache disorder | SNOMED | |
| 1076187 | Chronic headache due to and following craniotomy | SNOMED | |
| 37151048 | Chronic headache due to disorder | SNOMED | |
| 37171590 | Chronic headache due to disorder of temporomandibular joint | SNOMED | |
| 37172114 | Chronic headache due to infectious disease | SNOMED | |
| 37155929 | Chronic headache due to non-vascular intracranial disorder | SNOMED | |
| 4148541 | Chronic idiopathic anal pain | SNOMED | |
| 42539629 | Chronic idiopathic pain syndrome | SNOMED | |
| 43530652 | Chronic intractable migraine without aura | SNOMED | |
| 4168685 | Chronic intractable pain | SNOMED | |
| 4132891 | Chronic low back pain | SNOMED | |
| 37310730 | Chronic mechanical low back pain | SNOMED | |
| 42538794 | Chronic mechanical visceral pain | SNOMED | |
| 762769 | Chronic migraine without aura | SNOMED | |
| 765701 | Chronic migraine without aura, non-refractory | SNOMED | |
| 760968 | Chronic migraine without aura with status migrainosus | SNOMED | |
| 4318560 | Chronic mixed headache syndrome | SNOMED | |
| 42538688 | Chronic musculoskeletal pain | SNOMED | |
| 42538793 | Chronic musculoskeletal pain due to disease of nervous system | SNOMED | |
| 37171578 | Chronic musculoskeletal pain due to disorder | SNOMED | |
| 42538792 | Chronic musculoskeletal pain due to persistent inflammation | SNOMED | |
| 43530622 | Chronic neck pain | SNOMED | |
| 37016631 | Chronic neck pain for greater than 3 months | SNOMED | |
| 42536909 | Chronic neuropathic pain | SNOMED | |
| 37155930 | Chronic neuropathic pain following brain injury | SNOMED | |
| 37155931 | Chronic neuropathic pain following spinal cord injury | SNOMED | |
| 42536695 | Chronic nociceptive pain | SNOMED | |
| 43530731 | Chronic nonmalignant pain | SNOMED | |
| 4340938 | Chronic nonspecific abdominal pain | SNOMED | |
| 42538690 | Chronic orofacial pain | SNOMED | |
| 37171579 | Chronic orofacial pain due to disorder | SNOMED | |
| 37172119 | Chronic orofacial pain due to infectious disease | SNOMED | |
| 37155936 | Chronic orofacial pain due to nonvascular intracranial disorder | SNOMED | |
| 37171608 | Chronic orofacial pain due to temporomandibular joint disorder | SNOMED | |
| 436096 | Chronic pain | SNOMED | |
| 37312006 | Chronic pain after cancer treatment | SNOMED | |
| 4331953 | Chronic pain due to injury | SNOMED | |
| 42536908 | Chronic pain due to malignant neoplastic disease | SNOMED | |
| 37155938 | Chronic pain following amputation | SNOMED | |
| 37171604 | Chronic pain following arthroplasty | SNOMED | |
| 37151047 | Chronic pain following breast surgery | SNOMED | |
| 37171573 | Chronic pain following burn injury | SNOMED | |
| 37171605 | Chronic pain following cholecystectomy | SNOMED | |
| 37171575 | Chronic pain following hysterectomy | SNOMED | |
| 37108779 | Chronic pain following left total hip arthroplasty | SNOMED | |
| 37108780 | Chronic pain following left total knee arthroplasty | SNOMED | |
| 37171584 | Chronic pain following musculoskeletal injury | SNOMED | |
| 42539375 | Chronic pain following radiotherapy | SNOMED | |
| 37171574 | Chronic pain following repair of hernia | SNOMED | |
| 37108781 | Chronic pain following right total hip arthroplasty | SNOMED | |
| 37117142 | Chronic pain following right total knee arthroplasty | SNOMED | |
| 37171577 | Chronic pain following spinal surgery | SNOMED | |
| 42538790 | Chronic pain following surgical procedure for cancer | SNOMED | |
| 42538687 | Chronic pain following trauma | SNOMED | |
| 37155909 | Chronic pain following whiplash injury to neck | SNOMED | |
| 4105639 | Chronic painful neuropathy due to diabetes mellitus | SNOMED | |
| 37171585 | Chronic painful polyneuropathy | SNOMED | |
| 4044392 | Chronic painful polyneuropathy due to diabetes mellitus | SNOMED | |
| 42538791 | Chronic painful polyneuropathy following chemotherapy | SNOMED | |
| 37171586 | Chronic painful radiculopathy | SNOMED | |
| 43530631 | Chronic pain in coccyx for more than three months | SNOMED | |
| 4196899 | Chronic pain in face | SNOMED | |
| 43530684 | Chronic pain in male pelvis | SNOMED | |
| 37160602 | Chronic pain of cardiac implantable electronic device pocket following implantation of cardiac implantable electronic device | SNOMED | |
| 37108974 | Chronic pain of left foot | SNOMED | |
| 37108941 | Chronic pain of left upper limb | SNOMED | |
| 37108973 | Chronic pain of right foot | SNOMED | |
| 37108940 | Chronic pain of right upper limb | SNOMED | |
| 440704 | Chronic pain syndrome | SNOMED | |
| 442187 | Chronic paroxysmal hemicrania | SNOMED | |
| 4034006 | Chronic pelvic pain of female | SNOMED | |
| 4133035 | Chronic pelvic pain without obvious pathology | SNOMED | |
| 42538798 | Chronic peripheral neuropathic pain | SNOMED | |
| 37171583 | Chronic peripheral neuropathic pain following peripheral nerve injury | SNOMED | |
| 44783586 | Chronic post-concussion headache | SNOMED | |
| 43531612 | Chronic postoperative pain | SNOMED | |
| 44784644 | Chronic post-thoracotomy pain syndrome | SNOMED | |
| 377546 | Chronic post-traumatic headache | SNOMED | |
| 1246430 | Chronic primary cervical pain | SNOMED | |
| 42538786 | Chronic primary generalized pain | SNOMED | |
| 42538788 | Chronic primary headache | SNOMED | |
| 1246433 | Chronic primary low back pain | SNOMED | |
| 1246432 | Chronic primary musculoskeletal limb pain | SNOMED | |
| 42538787 | Chronic primary musculoskeletal pain | SNOMED | |
| 42538789 | Chronic primary orofacial pain | SNOMED | |
| 1246434 | Chronic primary thoracic pain | SNOMED | |
| 42538785 | Chronic primary visceral pain | SNOMED | |
| 37162891 | Chronic proctalgia | SNOMED | |
| 4138333 | Chronic prostatitis - chronic pelvic pain syndrome | SNOMED | |
| 40480423 | Chronic psychogenic pain | SNOMED | |
| 43530630 | Chronic sacral pain for greater than three months | SNOMED | |
| 37204166 | Chronic sacroiliac joint pain | SNOMED | |
| 4143863 | Chronic sciatica | SNOMED | |
| 42539376 | Chronic secondary facial pain | SNOMED | |
| 42538799 | Chronic secondary oral pain | SNOMED | |
| 608602 | Chronic shoulder pain | SNOMED | |
| 4168068 | Chronic sore throat | SNOMED | |
| 377853 | Chronic tension-type headache | SNOMED | |
| 43530661 | Chronic thoracic back pain | SNOMED | |
| 37209550 | Chronic urinary bladder pain | SNOMED | |
| 4142567 | Chronic vaginal pain | SNOMED | |
| 42537749 | Chronic visceral pain | SNOMED | |
| 37171580 | Chronic visceral pain due to disorder | SNOMED | |
| 42538796 | Chronic visceral pain due to persistent inflammation | SNOMED | |
| 42538795 | Chronic visceral pain due to vascular disorder | SNOMED | |
| 37171606 | Chronic visceral pain in abdominal region due to disorder | SNOMED | |
| 37171607 | Chronic visceral pain in pelvic region due to disorder | SNOMED | |
| 37155941 | Chronic visceral pain in thoracic region due to disorder | SNOMED | |
| 4134577 | Complex regional pain syndrome | SNOMED | |
| 42536258 | Complex regional pain syndrome of foot | SNOMED | |
| 42536260 | Complex regional pain syndrome of hand | SNOMED | |
| 42536259 | Complex regional pain syndrome of knee | SNOMED | |
| 42539474 | Complex regional pain syndrome of lower limb | SNOMED | |
| 46273207 | Complex regional pain syndrome of upper limb | SNOMED | |
| 761850 | Complex regional pain syndrome type 2 of bilateral lower limbs | SNOMED | |
| 765318 | Complex regional pain syndrome type 2 of bilateral upper limbs | SNOMED | |
| 42536233 | Complex regional pain syndrome type I | SNOMED | |
| 4256912 | Complex regional pain syndrome, type II | SNOMED | |
| 4237198 | Complex regional pain syndrome, type II, lower limb | SNOMED | |
| 42535432 | Complex regional pain syndrome type II of left lower limb | SNOMED | |
| 42535430 | Complex regional pain syndrome type II of left upper limb | SNOMED | |
| 42535433 | Complex regional pain syndrome type II of right lower limb | SNOMED | |
| 42535431 | Complex regional pain syndrome type II of right upper limb | SNOMED | |
| 4237315 | Complex regional pain syndrome, type II, upper limb | SNOMED | |
| 42535471 | Complex regional pain syndrome type I of bilateral lower limbs | SNOMED | |
| 42539169 | Complex regional pain syndrome type I of bilateral upper limbs | SNOMED | |
| 42535469 | Complex regional pain syndrome type I of left lower limb | SNOMED | |
| 42535470 | Complex regional pain syndrome type I of left upper limb | SNOMED | |
| 42539285 | Complex regional pain syndrome type I of right lower limb | SNOMED | |
| 42535472 | Complex regional pain syndrome type I of right upper limb | SNOMED | |
| 43530760 | Daily headache | SNOMED | |
| 601848 | Episodic migraine | SNOMED | |
| 42535475 | Episodic paroxysmal hemicrania | SNOMED | |
| 377545 | Episodic tension-type headache | SNOMED | |
| 40405599 | Fibromyalgia | SNOMED | |
| 36716799 | Frequent episodic tension-type headache | SNOMED | |
| 4010338 | Generalized chronic body pains | SNOMED | |
| 40480082 | Hemicrania continua | SNOMED | |
| 36716798 | Infrequent episodic tension-type headache | SNOMED | |
| 442774 | Intermittent claudication | SNOMED | |
| 37312529 | Intermittent claudication due to atherosclerosis of artery of limb | SNOMED | |
| 36717006 | Intermittent claudication of bilateral lower limbs co-occurrent and due to atherosclerosis | SNOMED | |
| 36717286 | Intermittent claudication of left lower limb co-occurrent and due to atherosclerosis | SNOMED | |
| 36712806 | Intermittent claudication of right lower limb co-occurrent and due to atherosclerosis | SNOMED | |
| 37016721 | Intermittent headache | SNOMED | |
| 4200298 | Intermittent pain | SNOMED | |
| 4133645 | Internal mammary artery syndrome | SNOMED | |
| 762083 | Intractable chronic headache following trauma | SNOMED | |
| 42535410 | Intractable chronic tension headache | SNOMED | |
| 762084 | Intractable episodic tension-type headache | SNOMED | |
| 46270414 | Intractable low back pain | SNOMED | |
| 4186213 | Jaw claudication | SNOMED | |
| 37118025 | Myofascial pain syndrome | SNOMED | |
| 4133038 | Myofascial pain syndrome of lower back | SNOMED | |
| 36684460 | Myofascial pain syndrome of lumbar spine | SNOMED | |
| 4150753 | Myofascial pain syndrome of neck | SNOMED | |
| 36684438 | Myofascial pain syndrome of thoracic spine | SNOMED | |
| 4133641 | Myofascial pain syndrome of thorax | SNOMED | |
| 4119307 | Neurogenic claudication | SNOMED | |
| 43530648 | New daily persistent headache | SNOMED | |
| 201347 | Pelvic congestion syndrome | SNOMED | |
| 42538612 | Persistent headache due to and following injury of head | SNOMED | |
| 37206118 | Persistent idiopathic facial pain | SNOMED | |
| 4150125 | Persistent pain following procedure | SNOMED | |
| 4345337 | Persistent prosthetic joint pain | SNOMED | |
| 4343237 | Persistent wound pain | SNOMED | |
| 37017894 | Post-mastectomy chronic pain syndrome | SNOMED | |
| 42539699 | Primary chronic pain | SNOMED | |
| 4316217 | Primary fibromyalgia syndrome | SNOMED | |
| 4161973 | Progressive angina | SNOMED | |
| 761867 | Reflex sympathetic dystrophy of bilateral upper limbs | SNOMED | |
| 4119942 | Stable angina | SNOMED | |
| 37209632 | Stable angina due to coronary arteriosclerosis | SNOMED | |
| 4141827 | Transformed migraine | SNOMED | |
| 4316222 | Venous intermittent claudication | SNOMED | |
| 1246661 | Vertebrogenic low back pain | SNOMED | |
| **Depression** | | | |
| 4031328 | Chronic major depressive disorder, single episode | SNOMED | |
| 4094358 | Chronic recurrent major depressive disorder | SNOMED | |
| 4269493 | Major depression in full remission | SNOMED | |
| 4148630 | Major depression in partial remission | SNOMED | |
| 4176002 | Major depression in remission | SNOMED | |
| 4154391 | Major depression, melancholic type | SNOMED | |
| 4282096 | Major depression, single episode | SNOMED | |
| 4323418 | Major depression single episode, in partial remission | SNOMED | |
| 37111697 | Major depression with psychotic features | SNOMED | |
| 4152280 | Major depressive disorder | SNOMED | |
| 45757195 | Major depressive disorder in mother complicating childbirth | SNOMED | |
| 45757196 | Major depressive disorder in mother complicating pregnancy | SNOMED | |
| 4181807 | Major depressive disorder, single episode with atypical features | SNOMED | |
| 4287238 | Major depressive disorder, single episode with catatonic features | SNOMED | |
| 4270907 | Major depressive disorder, single episode with melancholic features | SNOMED | |
| 4093584 | Major depressive disorder, single episode with postpartum onset | SNOMED | |
| 4336957 | Mild major depression | SNOMED | |
| 4195572 | Mild major depression, single episode | SNOMED | |
| 37109052 | Mild major depressive disorder co-occurrent with anxiety single episode | SNOMED | |
| 4228802 | Mild recurrent major depression | SNOMED | |
| 36715000 | Minimal major depression | SNOMED | |
| 36714999 | Minimal major depression single episode | SNOMED | |
| 36714997 | Minimal recurrent major depression | SNOMED | |
| 36714389 | Moderately severe major depression | SNOMED | |
| 36717389 | Moderately severe major depression single episode | SNOMED | |
| 36714998 | Moderately severe recurrent major depression | SNOMED | |
| 4307111 | Moderate major depression | SNOMED | |
| 4049623 | Moderate major depression, single episode | SNOMED | |
| 37109053 | Moderate major depressive disorder co-occurrent with anxiety single episode | SNOMED | |
| 4077577 | Moderate recurrent major depression | SNOMED | |
| 42534817 | Postpartum major depression in remission | SNOMED | |
| 4282316 | Recurrent major depression | SNOMED | |
| 4263748 | Recurrent major depression in full remission | SNOMED | |
| 4141454 | Recurrent major depression in partial remission | SNOMED | |
| 433991 | Recurrent major depression in remission | SNOMED | |
| 35615154 | Recurrent major depressive disorder co-occurrent with anxiety in full remission | SNOMED | |
| 35615155 | Recurrent major depressive disorder in partial remission co-occurrent with anxiety | SNOMED | |
| 4304140 | Recurrent major depressive disorder with atypical features | SNOMED | |
| 4220023 | Recurrent major depressive disorder with catatonic features | SNOMED | |
| 4205471 | Recurrent major depressive disorder with melancholic features | SNOMED | |
| 4324959 | Recurrent major depressive disorder with postpartum onset | SNOMED | |
| 432285 | Recurrent major depressive episodes | SNOMED | |
| 44805549 | Recurrent major depressive episodes, in partial remission | SNOMED | |
| 44813499 | Recurrent major depressive episodes, in remission | SNOMED | |
| 438998 | Recurrent major depressive episodes, mild | SNOMED | |
| 432883 | Recurrent major depressive episodes, moderate | SNOMED | |
| 44805542 | Recurrent major depressive episodes, severe | SNOMED | |
| 434911 | Recurrent major depressive episodes, severe, with psychosis | SNOMED | |
| 44805669 | Recurrent major depressive episodes, severe, with psychosis, psychosis in remission | SNOMED | |
| 35615151 | Recurrent mild major depressive disorder co-occurrent with anxiety | SNOMED | |
| 35615153 | Recurrent moderate major depressive disorder co-occurrent with anxiety | SNOMED | |
| 35615152 | Recurrent severe major depressive disorder co-occurrent with anxiety | SNOMED | |
| 42872722 | Severe major depression | SNOMED | |
| 42872411 | Severe major depression, single episode | SNOMED | |
| 441534 | Severe major depression, single episode, without psychotic features | SNOMED | |
| 438406 | Severe major depression, single episode, with psychotic features | SNOMED | |
| 4299785 | Severe major depression, single episode, with psychotic features, mood-congruent | SNOMED | |
| 4067409 | Severe major depression, single episode, with psychotic features, mood-incongruent | SNOMED | |
| 4327337 | Severe major depression without psychotic features | SNOMED | |
| 4250023 | Severe major depression with psychotic features | SNOMED | |
| 4144233 | Severe major depression with psychotic features, mood-congruent | SNOMED | |
| 4243822 | Severe major depression with psychotic features, mood-incongruent | SNOMED | |
| 37109054 | Severe major depressive disorder co-occurrent with anxiety single episode | SNOMED | |
| 43531624 | Severe recurrent major depression | SNOMED | |
| 435220 | Severe recurrent major depression without psychotic features | SNOMED | |
| 4154309 | Severe recurrent major depression with psychotic features | SNOMED | |
| 4141292 | Severe recurrent major depression with psychotic features, mood-congruent | SNOMED | |
| 4034842 | Severe recurrent major depression with psychotic features, mood-incongruent | SNOMED | |
| 4025677 | Single episode of major depression in full remission | SNOMED | |
| 44805550 | Single major depressive episode, in remission | SNOMED | |
| 439259 | Single major depressive episode, severe, with psychosis | SNOMED | |
| 44805668 | Single major depressive episode, severe, with psychosis, psychosis in remission | SNOMED | |
| **Bipolar Disorder** | | | |
| 439254 | Bipolar affective disorder, current episode depression | SNOMED | |
| 440078 | Bipolar affective disorder, current episode manic | SNOMED | |
| 435226 | Bipolar affective disorder, current episode mixed | SNOMED | |
| 439253 | Bipolar affective disorder, currently depressed, mild | SNOMED | |
| 437528 | Bipolar affective disorder, currently depressed, moderate | SNOMED | |
| 441834 | Bipolar affective disorder, currently manic, mild | SNOMED | |
| 433992 | Bipolar affective disorder, currently manic, moderate | SNOMED | |
| 439256 | Bipolar affective disorder, currently manic, severe, with psychosis | SNOMED | |
| 35624745 | Bipolar affective disorder, most recent episode mixed | SNOMED | |
| 436665 | Bipolar disorder | SNOMED | |
| 37209504 | Bipolar disorder caused by drug | SNOMED | |
| 4220618 | Bipolar disorder in full remission | SNOMED | |
| 436072 | Bipolar disorder in partial remission | SNOMED | |
| 4310821 | Bipolar disorder in remission | SNOMED | |
| 35624743 | Bipolar disorder, most recent episode depression | SNOMED | |
| 35624744 | Bipolar disorder, most recent episode manic | SNOMED | |
| 432876 | Bipolar I disorder | SNOMED | |
| 1076408 | Bipolar I disorder, current episode hypomanic | SNOMED | |
| 4107538 | Bipolar I disorder, most recent episode depressed with atypical features | SNOMED | |
| 4071442 | Bipolar I disorder, most recent episode depressed with catatonic features | SNOMED | |
| 4192865 | Bipolar I disorder, most recent episode depressed with melancholic features | SNOMED | |
| 4336405 | Bipolar I disorder, most recent episode depressed with postpartum onset | SNOMED | |
| 35624748 | Bipolar I disorder, most recent episode depression | SNOMED | |
| 4150985 | Bipolar I disorder, most recent episode hypomanic | SNOMED | |
| 35624747 | Bipolar I disorder, most recent episode manic | SNOMED | |
| 4073401 | Bipolar I disorder, most recent episode manic with catatonic features | SNOMED | |
| 4210024 | Bipolar I disorder, most recent episode manic with postpartum onset | SNOMED | |
| 4251178 | Bipolar I disorder, most recent episode mixed with catatonic features | SNOMED | |
| 4274957 | Bipolar I disorder, most recent episode mixed with postpartum onset | SNOMED | |
| 432866 | Bipolar I disorder, single manic episode | SNOMED | |
| 4148842 | Bipolar I disorder, single manic episode, in full remission | SNOMED | |
| 4301106 | Bipolar I disorder, single manic episode, in partial remission | SNOMED | |
| 4327669 | Bipolar I disorder, single manic episode, in remission | SNOMED | |
| 4338812 | Bipolar I disorder, single manic episode with catatonic features | SNOMED | |
| 4033390 | Bipolar I disorder, single manic episode with postpartum onset | SNOMED | |
| 4307956 | Bipolar II disorder | SNOMED | |
| 4172156 | Bipolar II disorder, most recent episode hypomanic | SNOMED | |
| 4037669 | Bipolar II disorder, most recent episode major depressive | SNOMED | |
| 4185096 | Bipolar II disorder, most recent episode major depressive with atypical features | SNOMED | |
| 4330846 | Bipolar II disorder, most recent episode major depressive with catatonic features | SNOMED | |
| 4144519 | Bipolar II disorder, most recent episode major depressive with melancholic features | SNOMED | |
| 4148934 | Bipolar II disorder, most recent episode major depressive with postpartum onset | SNOMED | |
| 37109940 | Bipolar type I disorder currently in full remission | SNOMED | |
| 37117177 | Bipolar type II disorder currently in full remission | SNOMED | |
| 4197669 | Chronic bipolar I disorder, most recent episode depressed | SNOMED | |
| 4001733 | Chronic bipolar II disorder, most recent episode major depressive | SNOMED | |
| 441836 | Depressed bipolar I disorder | SNOMED | |
| 435225 | Depressed bipolar I disorder in full remission | SNOMED | |
| 4177651 | Depressed bipolar I disorder in partial remission | SNOMED | |
| 4201739 | Depressed bipolar I disorder in remission | SNOMED | |
| 4287544 | Manic bipolar I disorder | SNOMED | |
| 436086 | Manic bipolar I disorder in full remission | SNOMED | |
| 442600 | Manic bipolar I disorder in partial remission | SNOMED | |
| 4166701 | Manic bipolar I disorder in remission | SNOMED | |
| 4028027 | Mild bipolar disorder | SNOMED | |
| 432290 | Mild bipolar I disorder, single manic episode | SNOMED | |
| 4324945 | Mild bipolar II disorder, most recent episode major depressive | SNOMED | |
| 437250 | Mild depressed bipolar I disorder | SNOMED | |
| 4215917 | Mild manic bipolar I disorder | SNOMED | |
| 440079 | Mild mixed bipolar I disorder | SNOMED | |
| 439250 | Mixed bipolar affective disorder | SNOMED | |
| 439245 | Mixed bipolar affective disorder, in full remission | SNOMED | |
| 44804961 | Mixed bipolar affective disorder, in partial remission | SNOMED | |
| 439249 | Mixed bipolar affective disorder, mild | SNOMED | |
| 439248 | Mixed bipolar affective disorder, moderate | SNOMED | |
| 44805540 | Mixed bipolar affective disorder, severe | SNOMED | |
| 439246 | Mixed bipolar affective disorder, severe, with psychosis | SNOMED | |
| 443906 | Mixed bipolar I disorder | SNOMED | |
| 4009648 | Mixed bipolar I disorder in full remission | SNOMED | |
| 437529 | Mixed bipolar I disorder in partial remission | SNOMED | |
| 433743 | Mixed bipolar I disorder in remission | SNOMED | |
| 4194222 | Moderate bipolar disorder | SNOMED | |
| 440067 | Moderate bipolar I disorder, single manic episode | SNOMED | |
| 4262111 | Moderate bipolar II disorder, most recent episode major depressive | SNOMED | |
| 4280361 | Moderate depressed bipolar I disorder | SNOMED | |
| 4307804 | Moderate manic bipolar I disorder | SNOMED | |
| 439785 | Moderate mixed bipolar I disorder | SNOMED | |
| 4333670 | Organic bipolar disorder | SNOMED | |
| 35622934 | Psychosis and severe depression co-occurrent and due to bipolar affective disorder | SNOMED | |
| 43021847 | Rapid cycling bipolar I disorder | SNOMED | |
| 37312578 | Rapid cycling bipolar II disorder | SNOMED | |
| 4244078 | Schizoaffective disorder, bipolar type | SNOMED | |
| 4155798 | Severe bipolar disorder | SNOMED | |
| 4200385 | Severe bipolar disorder without psychotic features | SNOMED | |
| 4195158 | Severe bipolar disorder with psychotic features | SNOMED | |
| 4322477 | Severe bipolar disorder with psychotic features, mood-congruent | SNOMED | |
| 4131027 | Severe bipolar disorder with psychotic features, mood-incongruent | SNOMED | |
| 4154283 | Severe bipolar I disorder | SNOMED | |
| 4030856 | Severe bipolar I disorder, single manic episode without psychotic features | SNOMED | |
| 4220617 | Severe bipolar I disorder, single manic episode with psychotic features | SNOMED | |
| 4030102 | Severe bipolar I disorder, single manic episode with psychotic features, mood-congruent | SNOMED | |
| 4312736 | Severe bipolar I disorder, single manic episode with psychotic features, mood-incongruent | SNOMED | |
| 4161200 | Severe bipolar II disorder | SNOMED | |
| 4045263 | Severe bipolar II disorder, most recent episode major depressive, in full remission | SNOMED | |
| 4283219 | Severe bipolar II disorder, most recent episode major depressive, in partial remission | SNOMED | |
| 4262272 | Severe bipolar II disorder, most recent episode major depressive, in remission | SNOMED | |
| 4217940 | Severe bipolar II disorder, most recent episode major depressive without psychotic features | SNOMED | |
| 4147991 | Severe bipolar II disorder, most recent episode major depressive with psychotic features | SNOMED | |
| 4000165 | Severe bipolar II disorder, most recent episode major depressive with psychotic features, mood-congruent | SNOMED | |
| 4051448 | Severe bipolar II disorder, most recent episode major depressive with psychotic features, mood-incongruent | SNOMED | |
| 42872413 | Severe depressed bipolar I disorder | SNOMED | |
| 442570 | Severe depressed bipolar I disorder without psychotic features | SNOMED | |
| 436386 | Severe depressed bipolar I disorder with psychotic features | SNOMED | |
| 4182998 | Severe depressed bipolar I disorder with psychotic features, mood-congruent | SNOMED | |
| 4094507 | Severe depressed bipolar I disorder with psychotic features, mood-incongruent | SNOMED | |
| 43020451 | Severe manic bipolar I disorder | SNOMED | |
| 443797 | Severe manic bipolar I disorder without psychotic features | SNOMED | |
| 4102603 | Severe manic bipolar I disorder with psychotic features | SNOMED | |
| 438129 | Severe manic bipolar I disorder with psychotic features, mood-congruent | SNOMED | |
| 4141603 | Severe manic bipolar I disorder with psychotic features, mood-incongruent | SNOMED | |
| 42872412 | Severe mixed bipolar I disorder | SNOMED | |
| 372599 | Severe mixed bipolar I disorder without psychotic features | SNOMED | |
| 439001 | Severe mixed bipolar I disorder with psychotic features | SNOMED | |
| 4276670 | Severe mixed bipolar I disorder with psychotic features, mood-congruent | SNOMED | |
| 4031928 | Severe mixed bipolar I disorder with psychotic features, mood-incongruent | SNOMED | |
| **Schizophrenia** | | | |
| 436071 | Acute exacerbation of chronic catatonic schizophrenia | SNOMED | |
| 437243 | Acute exacerbation of chronic paranoid schizophrenia | SNOMED | |
| 4100366 | Acute exacerbation of chronic schizophrenia | SNOMED | |
| 439275 | Acute exacerbation of subchronic catatonic schizophrenia | SNOMED | |
| 434901 | Acute exacerbation of subchronic paranoid schizophrenia | SNOMED | |
| 432864 | Acute schizophrenic episode | SNOMED | |
| 35610094 | Cataleptic schizophrenia | SNOMED | |
| 433996 | Catatonic schizophrenia | SNOMED | |
| 434332 | Catatonic schizophrenia in remission | SNOMED | |
| 4102660 | Cenesthopathic schizophrenia | SNOMED | |
| 1076115 | Childhood-onset schizophrenia | SNOMED | |
| 441538 | Chronic catatonic schizophrenia | SNOMED | |
| 440368 | Chronic disorganized schizophrenia | SNOMED | |
| 436385 | Chronic disorganized schizophrenia with acute exacerbation | SNOMED | |
| 436944 | Chronic paranoid schizophrenia | SNOMED | |
| 436673 | Chronic residual schizophrenia | SNOMED | |
| 435218 | Chronic residual schizophrenia with acute exacerbations | SNOMED | |
| 435782 | Chronic schizophrenia | SNOMED | |
| 4105330 | Chronic undifferentiated schizophrenia | SNOMED | |
| 4194671 | Chronic undifferentiated schizophrenia with acute exacerbations | SNOMED | |
| 441828 | Disorganized schizophrenia | SNOMED | |
| 436947 | Disorganized schizophrenia in remission | SNOMED | |
| 37163122 | Early onset schizophrenia | SNOMED | |
| 4244059 | Involutional paraphrenia | SNOMED | |
| 432300 | Latent schizophrenia in remission | SNOMED | |
| 4219539 | Late onset schizophrenia | SNOMED | |
| 40480879 | Lethal catatonia | SNOMED | |
| 433450 | Paranoid schizophrenia | SNOMED | |
| 435217 | Paranoid schizophrenia in remission | SNOMED | |
| 434318 | Paraphrenia | SNOMED | |
| 439004 | Residual schizophrenia | SNOMED | |
| 440686 | Residual schizophrenia in remission | SNOMED | |
| 434321 | Schizoaffective schizophrenia in remission | SNOMED | |
| 435783 | Schizophrenia | SNOMED | |
| 435219 | Schizophrenia in remission | SNOMED | |
| 4100365 | Schizophrenic disorders | SNOMED | |
| 4085662 | Schizophrenic prodrome | SNOMED | |
| 436067 | Simple schizophrenia | SNOMED | |
| 433990 | Subchronic catatonic schizophrenia | SNOMED | |
| 438724 | Subchronic disorganized schizophrenia | SNOMED | |
| 436384 | Subchronic disorganized schizophrenia with acute exacerbations | SNOMED | |
| 432299 | Subchronic paranoid schizophrenia | SNOMED | |
| 433442 | Subchronic residual schizophrenia | SNOMED | |
| 444396 | Subchronic residual schizophrenia with acute exacerbations | SNOMED | |
| 440373 | Subchronic schizophrenia | SNOMED | |
| 432598 | Subchronic schizophrenia with acute exacerbations | SNOMED | |
| 4310121 | Subchronic undifferentiated schizophrenia | SNOMED | |
| 4321694 | Subchronic undifferentiated schizophrenia with acute exacerbations | SNOMED | |
| 4008566 | Undifferentiated schizophrenia | SNOMED | |
| 4213979 | Undifferentiated schizophrenia in remission | SNOMED | |
| **Cannabis Use Disorder** | | | |
| 434327 | Cannabis abuse | SNOMED | |
| 440387 | Cannabis dependence | SNOMED | |
| 433452 | Cannabis dependence, continuous | SNOMED | |
| 437838 | Cannabis dependence, episodic | SNOMED | |
| 440996 | Cannabis dependence in remission | SNOMED | |
| 4103419 | Nondependent cannabis abuse | SNOMED | |
| 434019 | Nondependent cannabis abuse, continuous | SNOMED | |
| 434328 | Nondependent cannabis abuse, episodic | SNOMED | |
| 435231 | Nondependent cannabis abuse in remission | SNOMED | |
| **Alcohol Use Disorder** | | | |
| 4338024 | Absinthe addiction | SNOMED | |
| 433753 | Alcohol abuse | SNOMED | |
| 435243 | Alcohol dependence | SNOMED | |
| 45757131 | Alcohol dependence in childbirth | SNOMED | |
| 45757093 | Alcohol dependence in pregnancy | SNOMED | |
| 4218106 | Alcoholism | SNOMED | |
| 439005 | Chronic alcoholism in remission | SNOMED | |
| 436953 | Continuous chronic alcoholism | SNOMED | |
| 435532 | Episodic chronic alcoholism | SNOMED | |
| 37017329 | Mild alcohol dependence | SNOMED | |
| 37018356 | Moderate alcohol dependence | SNOMED | |
| 4152165 | Nondependent alcohol abuse | SNOMED | |
| 435534 | Nondependent alcohol abuse, continuous | SNOMED | |
| 440685 | Nondependent alcohol abuse, episodic | SNOMED | |
| 441276 | Nondependent alcohol abuse in remission | SNOMED | |
| 4109691 | Persistent alcohol abuse | SNOMED | |
| 37017563 | Severe alcohol dependence | SNOMED | |
| **Tobacco Use Disorder** | | | |
| 765685 | Continuous dependence on snuff use | SNOMED | |
| 764471 | Episodic dependence on chewing tobacco | SNOMED | |
| 764469 | Episodic dependence on cigarette smoking | SNOMED | |
| 764470 | Episodic dependence on snuff use | SNOMED | |
| 37169544 | Tobacco dependence caused by chewing tobacco in remission | SNOMED | |
| 4099811 | Tobacco dependence, continuous | SNOMED | |
| 4103417 | Tobacco dependence, episodic | SNOMED | |
| 4103418 | Tobacco dependence in remission | SNOMED | |
| 437264 | Tobacco dependence syndrome | SNOMED | |
| **Human Immunodeficiency Virus** | | | |
| 37017285 | Acquired hemolytic anemia co-occurrent with human immunodeficiency virus infection | SNOMED | |
| 45757102 | Acquired immune deficiency syndrome complicating childbirth | SNOMED | |
| 4128060 | Acquired immune deficiency syndrome-related nephropathy | SNOMED | |
| 4221489 | Acquired immunodeficiency syndrome-associated disorder | SNOMED | |
| 37017266 | Acute endocarditis co-occurrent with human immunodeficiency virus infection | SNOMED | |
| 4222405 | Acute endocarditis with AIDS (acquired immunodeficiency syndrome) | SNOMED | |
| 4008081 | Acute HIV infection | SNOMED | |
| 37017282 | Agranulocytosis co-occurrent with human immunodeficiency virus infection | SNOMED | |
| 4224553 | Agranulocytosis with AIDS (acquired immunodeficiency syndrome) | SNOMED | |
| 4267414 | AIDS | SNOMED | |
| 37166798 | AIDS (acquired immunodeficiency syndrome) wasting syndrome | SNOMED | |
| 37017132 | Anemia co-occurrent with human immunodeficiency virus infection | SNOMED | |
| 37019055 | Aplastic anemia co-occurrent with human immunodeficiency virus infection | SNOMED | |
| 4225810 | Aplastic anemia with AIDS (acquired immunodeficiency syndrome) | SNOMED | |
| 4201627 | Aseptic meningitis due to human immunodeficiency virus infection | SNOMED | |
| 37017456 | Aspergillosis co-occurrent with human immunodeficiency virus infection | SNOMED | |
| 42538959 | Asymptomatic human immunodeficiency virus A1 infection | SNOMED | |
| 42538960 | Asymptomatic human immunodeficiency virus A2 infection | SNOMED | |
| 4241530 | Asymptomatic human immunodeficiency virus infection | SNOMED | |
| 4239722 | Asymptomatic human immunodeficiency virus infection in pregnancy | SNOMED | |
| 37019058 | Bacterial pneumonia co-occurrent with human immunodeficiency virus infection | SNOMED | |
| 4223032 | Bacterial pneumonia with AIDS (acquired immunodeficiency syndrome) | SNOMED | |
| 4226790 | Blindness with AIDS (acquired immunodeficiency syndrome) | SNOMED | |
| 37017595 | Burkitt lymphoma co-occurrent with human immunodeficiency virus infection | SNOMED | |
| 37017092 | Candidiasis of esophagus co-occurrent with human immunodeficiency virus infection | SNOMED | |
| 4222062 | Candidiasis of lung with AIDS (acquired immunodeficiency syndrome) | SNOMED | |
| 37017254 | Candidiasis of mouth co-occurrent with human immunodeficiency virus infection | SNOMED | |
| 4226792 | Candidiasis of mouth with AIDS (acquired immunodeficiency syndrome) | SNOMED | |
| 36714339 | Candidiasis of upper respiratory tract co-occurrent with human immunodeficiency virus infection | SNOMED | |
| 37017276 | Cardiomyopathy co-occurrent with human immunodeficiency virus infection | SNOMED | |
| 4226110 | Central nervous disorder with AIDS (acquired immunodeficiency syndrome) | SNOMED | |
| 4223276 | Central nervous system demyelinating disease with AIDS (acquired immunodeficiency syndrome) | SNOMED | |
| 44783780 | Cholangitis with AIDS (acquired immunodeficiency syndrome) | SNOMED | |
| 3654682 | Chronic hepatitis C co-occurrent with human immunodeficiency virus infection | SNOMED | |
| 37017210 | Chronic infection caused by herpes simplex virus co-occurrent with human immunodeficiency virus infection | SNOMED | |
| 4224740 | Coccidioidomycosis with AIDS (acquired immunodeficiency syndrome) | SNOMED | |
| 37017453 | Coccidiosis co-occurrent with human immunodeficiency virus infection | SNOMED | |
| 4171124 | Congenital acquired immune deficiency syndrome | SNOMED | |
| 4180254 | Congenital human immunodeficiency virus infection | SNOMED | |
| 4171125 | Congenital human immunodeficiency virus positive status syndrome | SNOMED | |
| 4222712 | Cryptococcosis with AIDS (acquired immunodeficiency syndrome) | SNOMED | |
| 37017549 | Dementia co-occurrent with human immunodeficiency virus infection | SNOMED | |
| 4228133 | Dementia with AIDS (acquired immunodeficiency syndrome) | SNOMED | |
| 37017294 | Demyelinating disease of central nervous system co-occurrent with human immunodeficiency virus infection | SNOMED | |
| 4227229 | Dermatomycosis with AIDS (acquired immunodeficiency syndrome) | SNOMED | |
| 37018935 | Dermatophytosis co-occurrent with human immunodeficiency virus infection | SNOMED | |
| 37017442 | Diffuse non-Hodgkin immunoblastic lymphoma co-occurrent with human immunodeficiency virus infection | SNOMED | |
| 37017319 | Disorder of central nervous system co-occurrent with human immunodeficiency virus infection | SNOMED | |
| 37018711 | Disorder of eye proper co-occurrent with human immunodeficiency virus infection | SNOMED | |
| 37017094 | Disorder of gastrointestinal tract co-occurrent with human immunodeficiency virus infection | SNOMED | |
| 37017260 | Disorder of kidney co-occurrent with human immunodeficiency virus infection | SNOMED | |
| 37017279 | Disorder of peripheral nervous system co-occurrent with human immunodeficiency virus infection | SNOMED | |
| 37017244 | Disorder of respiratory system co-occurrent with human immunodeficiency virus infection | SNOMED | |
| 37017125 | Disorder of skin co-occurrent with human immunodeficiency virus infection | SNOMED | |
| 37017259 | Disorder of spinal cord co-occurrent with human immunodeficiency virus infection | SNOMED | |
| 37017655 | Disseminated atypical infection caused by Mycobacterium co-occurrent with human immunodeficiency virus infection | SNOMED | |
| 4227380 | Disseminated candidiasis with AIDS (acquired immunodeficiency syndrome) | SNOMED | |
| 37019052 | Disseminated infection caused by Strongyloides co-occurrent with human immunodeficiency virus infection | SNOMED | |
| 4228754 | Dyspnea with AIDS (acquired immunodeficiency syndrome) | SNOMED | |
| 3654950 | Encephalitis caused by human immunodeficiency virus type 1 | SNOMED | |
| 3654900 | Encephalitis caused by human immunodeficiency virus type 2 | SNOMED | |
| 4222250 | Encephalitis with AIDS (acquired immunodeficiency syndrome) | SNOMED | |
| 4228612 | Encephalomyelitis with AIDS (acquired immunodeficiency syndrome) | SNOMED | |
| 4228440 | Encephalopathy with AIDS (acquired immunodeficiency syndrome) | SNOMED | |
| 37017265 | Enlargement of liver co-occurrent with human immunodeficiency virus infection | SNOMED | |
| 37017106 | Eruption of skin co-occurrent with human immunodeficiency virus infection | SNOMED | |
| 37116831 | Extrapulmonary tuberculosis co-occurrent with human immunodeficiency virus infection | SNOMED | |
| 4228272 | Failure to thrive in infant with AIDS (acquired immunodeficiency syndrome) | SNOMED | |
| 4221911 | Fatigue with AIDS (acquired immunodeficiency syndrome) | SNOMED | |
| 37017586 | Focal segmental glomerulosclerosis co-occurrent with human immunodeficiency virus infection | SNOMED | |
| 37017261 | Gastrointestinal malabsorption syndrome co-occurrent with human immunodeficiency virus infection | SNOMED | |
| 37017093 | Heart disease co-occurrent with human immunodeficiency virus infection | SNOMED | |
| 4222402 | Hematopoietic system disease with AIDS (acquired immunodeficiency syndrome) | SNOMED | |
| 37017209 | Hemophagocytic syndrome with human immunodeficiency virus infection | SNOMED | |
| 4222224 | Hepatomegaly with AIDS (acquired immunodeficiency syndrome) | SNOMED | |
| 4227539 | Herpes zoster with AIDS (acquired immunodeficiency syndrome) | SNOMED | |
| 4225170 | Histoplasmosis with AIDS (acquired immunodeficiency syndrome) | SNOMED | |
| 46284256 | HIV (human immunodeficiency virus) disease resulting in haematological and immunological abnormalities | SNOMED | |
| 4314426 | HIV infection with acute lymphadenitis | SNOMED | |
| 4172009 | HIV infection with infectious mononucleosis-like syndrome | SNOMED | |
| 36715476 | Human immunodeficiency virus complicating pregnancy childbirth and the puerperium | SNOMED | |
| 4161950 | Human immunodeficiency virus encephalitis | SNOMED | |
| 4262297 | Human immunodeficiency virus encephalopathy | SNOMED | |
| 4340791 | Human immunodeficiency virus enteropathy | SNOMED | |
| 432554 | Human immunodeficiency virus II infection | SNOMED | |
| 4253472 | Human immunodeficiency virus I infection | SNOMED | |
| 439727 | Human immunodeficiency virus infection | SNOMED | |
| 4087603 | Human immunodeficiency virus infection constitutional disease | SNOMED | |
| 36687122 | Human immunodeficiency virus infection with cognitive impairment | SNOMED | |
| 4087604 | Human immunodeficiency virus infection with neurological disease | SNOMED | |
| 4092686 | Human immunodeficiency virus infection with secondary clinical infectious disease | SNOMED | |
| 45757132 | Human immunodeficiency virus in mother complicating childbirth | SNOMED | |
| 4047624 | Human immunodeficiency virus leukoencephalopathy | SNOMED | |
| 606047 | Human immunodeficiency virus modified skin disease | SNOMED | |
| 4347288 | Human immunodeficiency virus myopathy | SNOMED | |
| 42536591 | Human immunodeficiency virus World Health Organization 2007 stage 1 co-occurrent with malaria | SNOMED | |
| 42536590 | Human immunodeficiency virus World Health Organization 2007 stage 1 co-occurrent with tuberculosis | SNOMED | |
| 42536593 | Human immunodeficiency virus World Health Organization 2007 stage 2 co-occurrent with malaria | SNOMED | |
| 42536592 | Human immunodeficiency virus World Health Organization 2007 stage 2 co-occurrent with tuberculosis | SNOMED | |
| 42536594 | Human immunodeficiency virus World Health Organization 2007 stage 3 co-occurrent with malaria | SNOMED | |
| 42539031 | Human immunodeficiency virus World Health Organization 2007 stage 3 co-occurrent with tuberculosis | SNOMED | |
| 42536596 | Human immunodeficiency virus World Health Organization 2007 stage 4 co-occurrent with malaria | SNOMED | |
| 42536595 | Human immunodeficiency virus World Health Organization 2007 stage 4 co-occurrent with tuberculosis | SNOMED | |
| 4228760 | Hyperhidrosis with AIDS (acquired immunodeficiency syndrome) | SNOMED | |
| 37018063 | Immune reconstitution inflammatory syndrome caused by human immunodeficiency virus infection | SNOMED | |
| 36674252 | Infection caused by Coccidia co-occurrent with acquired immunodeficiency syndrome | SNOMED | |
| 37017550 | Infection caused by Cryptosporidium co-occurrent with human immunodeficiency virus infection | SNOMED | |
| 37017446 | Infection caused by Cytomegalovirus co-occurrent with human immunodeficiency virus infection | SNOMED | |
| 37017454 | Infection caused by herpes simplex virus co-occurrent with human immunodeficiency virus infection | SNOMED | |
| 37017457 | Infection caused by herpes zoster virus co-occurrent with human immunodeficiency virus infection | SNOMED | |
| 36674254 | Infection caused by Isospora co-occurrent with acquired immunodeficiency syndrome | SNOMED | |
| 37017295 | Infection caused by Nocardia co-occurrent with human immunodeficiency virus infection | SNOMED | |
| 37017248 | Infection caused by Pneumocystis co-occurrent with human immunodeficiency virus infection | SNOMED | |
| 37019042 | Infection caused by Salmonella co-occurrent with human immunodeficiency virus infection | SNOMED | |
| 37017124 | Infection caused by Strongyloides co-occurrent with human immunodeficiency virus infection | SNOMED | |
| 36674253 | Infection caused by Toxoplasma gondii co-occurrent with acquired immunodeficiency syndrome | SNOMED | |
| 40493262 | Infectious disease with acquired immune deficiency syndrome | SNOMED | |
| 37017318 | Infectious gastroenteritis co-occurrent with human immunodeficiency virus infection | SNOMED | |
| 4225989 | Infectious gastroenteritis with AIDS (acquired immunodeficiency syndrome) | SNOMED | |
| 37162220 | Infectious mononucleosis caused by human immunodeficiency virus | SNOMED | |
| 37162221 | Infectious mononucleosis caused by human immunodeficiency virus type I | SNOMED | |
| 37017284 | Infective arthritis co-occurrent with human immunodeficiency virus infection | SNOMED | |
| 4228290 | Infective arthritis with AIDS (acquired immunodeficiency syndrome) | SNOMED | |
| 4222383 | Intestinal malabsorption with AIDS (acquired immunodeficiency syndrome) | SNOMED | |
| 37116830 | Invasive carcinoma of uterine cervix co-occurrent with human immunodeficiency virus infection | SNOMED | |
| 37017296 | Isosporiasis co-occurrent with human immunodeficiency virus infection | SNOMED | |
| 4224096 | Kaposi's sarcoma with AIDS (acquired immunodeficiency syndrome) | SNOMED | |
| 4228757 | Low vision with AIDS (acquired immunodeficiency syndrome) | SNOMED | |
| 37017263 | Lymphadenopathy co-occurrent with human immunodeficiency virus infection | SNOMED | |
| 4225027 | Malaise with AIDS (acquired immunodeficiency syndrome) | SNOMED | |
| 4226357 | Malignant neoplasm with AIDS (acquired immunodeficiency syndrome) | SNOMED | |
| 37017320 | Malignant neoplastic disease co-occurrent with human immunodeficiency virus infection | SNOMED | |
| 4225193 | Microsporidiosis associated with acquired immunodeficiency syndrome | SNOMED | |
| 37017580 | Microsporidiosis co-occurrent with human immunodeficiency virus infection | SNOMED | |
| 37017652 | Multidermatomal infection caused by Herpes zoster co-occurrent with human immunodeficiency virus infection | SNOMED | |
| 4221607 | Mycobacteriosis with AIDS (acquired immunodeficiency syndrome) | SNOMED | |
| 37017126 | Myelitis co-occurrent with human immunodeficiency virus infection | SNOMED | |
| 4227974 | Myelitis with AIDS (acquired immunodeficiency syndrome) | SNOMED | |
| 4224078 | Myelopathy with AIDS (acquired immunodeficiency syndrome) | SNOMED | |
| 37017108 | Myocarditis co-occurrent with human immunodeficiency virus infection | SNOMED | |
| 4227053 | Myocarditis with AIDS (acquired immunodeficiency syndrome) | SNOMED | |
| 37017424 | Nephrotic syndrome co-occurrent with human immunodeficiency virus infection | SNOMED | |
| 37019034 | Neuralgia co-occurrent with human immunodeficiency virus infection | SNOMED | |
| 4228745 | Neuralgia with AIDS (acquired immunodeficiency syndrome) | SNOMED | |
| 37017262 | Neuritis co-occurrent with human immunodeficiency virus infection | SNOMED | |
| 4227519 | Neuritis with AIDS (acquired immunodeficiency syndrome) | SNOMED | |
| 4048033 | Neuropathy caused by human immunodeficiency virus | SNOMED | |
| 4211956 | Neutropenia with AIDS (acquired immunodeficiency syndrome) | SNOMED | |
| 4224578 | Nocardiosis with AIDS (acquired immunodeficiency syndrome) | SNOMED | |
| 4223738 | Noninfectious gastroenteritis with AIDS (acquired immunodeficiency syndrome) | SNOMED | |
| 4227077 | Nutritional deficiency with AIDS (acquired immunodeficiency syndrome) | SNOMED | |
| 37017579 | Opportunistic mycosis co-occurrent with human immunodeficiency virus infection | SNOMED | |
| 37017249 | Organic brain syndrome co-occurrent with human immunodeficiency virus infection | SNOMED | |
| 4227205 | Organic brain syndrome with AIDS (acquired immunodeficiency syndrome) | SNOMED | |
| 4224860 | Organic dementia with AIDS (acquired immunodeficiency syndrome) | SNOMED | |
| 36716524 | Parkinsonism due to human immunodeficiency virus infection | SNOMED | |
| 4236860 | Pediatric human immunodeficiency virus infection | SNOMED | |
| 4320032 | Persistent generalized lymphadenopathy | SNOMED | |
| 4221503 | Pneumococcal pneumonia with AIDS (acquired immunodeficiency syndrome) | SNOMED | |
| 4226502 | Pneumocystosis with AIDS (acquired immunodeficiency syndrome) | SNOMED | |
| 4228277 | Pneumonia with AIDS (acquired immunodeficiency syndrome) | SNOMED | |
| 37018721 | Polyneuropathy co-occurrent with human immunodeficiency virus infection | SNOMED | |
| 4226662 | Polyneuropathy with AIDS (acquired immunodeficiency syndrome) | SNOMED | |
| 37017247 | Presenile dementia co-occurrent with human immunodeficiency virus infection | SNOMED | |
| 4224240 | Presenile dementia with AIDS (acquired immunodeficiency syndrome) | SNOMED | |
| 37017112 | Primary cerebral lymphoma co-occurrent with human immunodeficiency virus infection | SNOMED | |
| 606040 | Primary human immunodeficiency virus infection | SNOMED | |
| 4227969 | Primary lymphoma of brain with AIDS (acquired immunodeficiency syndrome) | SNOMED | |
| 37017246 | Progressive multifocal leukoencephalopathy co-occurrent with human immunodeficiency virus infection | SNOMED | |
| 4226631 | Progressive multifocal leukoencephalopathy with AIDS (acquired immunodeficiency syndrome) | SNOMED | |
| 37017455 | Pyrexia of unknown origin co-occurrent with human immunodeficiency virus infection | SNOMED | |
| 37018714 | Radiculitis co-occurrent with human immunodeficiency virus infection | SNOMED | |
| 4227072 | Radiculitis with AIDS (acquired immunodeficiency syndrome) | SNOMED | |
| 37017278 | Recurrent bacterial pneumonia co-occurrent with human immunodeficiency virus infection | SNOMED | |
| 37018755 | Recurrent salmonella sepsis co-occurrent with human immunodeficiency virus infection | SNOMED | |
| 37017425 | Renal failure syndrome co-occurrent with human immunodeficiency virus infection | SNOMED | |
| 4225643 | Respiratory disorder with AIDS (acquired immunodeficiency syndrome) | SNOMED | |
| 37017243 | Reticulosarcoma co-occurrent with human immunodeficiency virus infection | SNOMED | |
| 4226388 | Reticulosarcoma with AIDS (acquired immunodeficiency syndrome) | SNOMED | |
| 4223918 | Retinal vascular changes with AIDS (acquired immunodeficiency syndrome) | SNOMED | |
| 4228115 | Retinopathy with AIDS (acquired immunodeficiency syndrome) | SNOMED | |
| 4226629 | Salivary gland disease with AIDS (acquired immunodeficiency syndrome) | SNOMED | |
| 4225017 | Salmonella infection with AIDS (acquired immunodeficiency syndrome) | SNOMED | |
| 44784137 | Sepsis with acquired immunodeficiency syndrome | SNOMED | |
| 4224566 | Skin disorder with AIDS (acquired immunodeficiency syndrome) | SNOMED | |
| 4220603 | Skin rash with AIDS (acquired immunodeficiency syndrome) | SNOMED | |
| 37017082 | Splenomegaly co-occurrent with human immunodeficiency virus infection | SNOMED | |
| 4223597 | Strongyloidiasis with AIDS (acquired immunodeficiency syndrome) | SNOMED | |
| 37017071 | Subacute adenoviral encephalitis co-occurrent with human immunodeficiency virus infection | SNOMED | |
| 4222717 | Subacute adenoviral encephalitis with AIDS (acquired immunodeficiency syndrome) | SNOMED | |
| 4227350 | Subacute endocarditis with AIDS (acquired immunodeficiency syndrome) | SNOMED | |
| 4224425 | Subacute myocarditis with AIDS (acquired immunodeficiency syndrome) | SNOMED | |
| 45769864 | Symptomatic human immunodeficiency virus (HIV) I infection | SNOMED | |
| 43531586 | Symptomatic human immunodeficiency virus infection | SNOMED | |
| 4226905 | Thrombocytopenia with AIDS (acquired immunodeficiency syndrome) | SNOMED | |
| 4226658 | Tuberculosis with AIDS (acquired immunodeficiency syndrome) | SNOMED | |
| 4045982 | Vacuolar myelopathy | SNOMED | |
| 4225318 | Viral pneumonia with AIDS (acquired immunodeficiency syndrome) | SNOMED | |
| 37017283 | Visual impairment co-occurrent with human immunodeficiency virus infection | SNOMED | |
| **Hepatitis C Virus** | | | |
| 192242 | Acute hepatitis C | SNOMED | |
| 45769525 | Chronic active hepatitis C | SNOMED | |
| 3654685 | Chronic hepatitis B co-occurrent with hepatitis C and hepatitis D | SNOMED | |
| 198964 | Chronic hepatitis C | SNOMED | |
| 35625141 | Chronic hepatitis C caused by Hepatitis C virus genotype 1 | SNOMED | |
| 35625296 | Chronic hepatitis C caused by Hepatitis C virus genotype 1a | SNOMED | |
| 35625295 | Chronic hepatitis C caused by Hepatitis C virus genotype 1b | SNOMED | |
| 35625139 | Chronic hepatitis C caused by Hepatitis C virus genotype 2 | SNOMED | |
| 35625040 | Chronic hepatitis C caused by Hepatitis C virus genotype 3 | SNOMED | |
| 35625140 | Chronic hepatitis C caused by Hepatitis C virus genotype 4 | SNOMED | |
| 35624867 | Chronic hepatitis C caused by hepatitis C virus genotype 5 | SNOMED | |
| 35624866 | Chronic hepatitis C caused by hepatitis C virus genotype 6 | SNOMED | |
| 3654682 | Chronic hepatitis C co-occurrent with human immunodeficiency virus infection | SNOMED | |
| 45766656 | Chronic hepatitis C with stage 2 fibrosis | SNOMED | |
| 45757726 | Chronic hepatitis C with stage 3 fibrosis | SNOMED | |
| 763021 | Chronic viral hepatitis C with hepatic coma | SNOMED | |
| 40483136 | Hepatitis B and hepatitis C | SNOMED | |
| 44809233 | Hepatitis C genotype 1 | SNOMED | |
| 44809234 | Hepatitis C genotype 2 | SNOMED | |
| 44809236 | Hepatitis C genotype 3 | SNOMED | |
| 44809237 | Hepatitis C genotype 4 | SNOMED | |
| 44809238 | Hepatitis C genotype 5 | SNOMED | |
| 44809239 | Hepatitis C genotype 6 | SNOMED | |
| 37172860 | Perinatal hepatitis C | SNOMED | |
| 45773146 | Reactivation of hepatitis C viral hepatitis | SNOMED | |
| 37162668 | Recurrent hepatitis C virus induced liver disease following liver transplant | SNOMED | |
| 197494 | Viral hepatitis C | SNOMED | |
| 37151819 | Viral hepatitis C in mother during pregnancy | SNOMED | |
| **Hepatitis B Virus** | | | |
| 4173584 | Chronic active type B viral hepatitis | SNOMED | |
| 4009793 | Chronic aggressive type B viral hepatitis | SNOMED | |
| 3654685 | Chronic hepatitis B co-occurrent with hepatitis C and hepatitis D | SNOMED | |
| 37175349 | Chronic hepatitis B during pregnancy | SNOMED | |
| 4296554 | Chronic persistent type B viral hepatitis | SNOMED | |
| 194574 | Chronic type B viral hepatitis | SNOMED | |
| 192240 | Chronic viral hepatitis B with hepatitis D | SNOMED | |
| 439674 | Chronic viral hepatitis B without delta-agent | SNOMED | |
| 197493 | Hepatitis D superinfection of hepatitis B carrier | SNOMED | |
| 37017654 | Occult chronic type B viral hepatitis | SNOMED | |

### **Table S4:** RxNorm vocabulary codes for medications for opioid use disorder.

Listed are concept IDs and names for methadone, buprenorphine, and naltrexone products.

| **Concept Id** | **Concept Name** | **Vocabulary** |
| --- | --- | --- |
| **Methadone** | | |
| 793092 | Diskets Oral Liquid Product | RxNorm |
| 793093 | Diskets Oral Product | RxNorm |
| 36234934 | Dolophine Injectable Product | RxNorm |
| 36234935 | Dolophine Oral Product | RxNorm |
| 36234936 | Dolophine Pill | RxNorm |
| 36232599 | Martindale Methadone DTF Oral Liquid Product | RxNorm |
| 36232600 | Martindale Methadone DTF Oral Product | RxNorm |
| 1103640 | methadone | RxNorm |
| 19112776 | methadone 45 MG/ML | RxNorm |
| 19112777 | methadone 45 MG/ML Oral Solution | RxNorm |
| 40166290 | methadone hydrochloride 0.25 MG/ML | RxNorm |
| 40166291 | methadone hydrochloride 0.25 MG/ML Oral Solution | RxNorm |
| 40166299 | methadone hydrochloride 0.4 MG/ML | RxNorm |
| 40166300 | methadone hydrochloride 0.4 MG/ML Oral Solution | RxNorm |
| 40166303 | methadone hydrochloride 0.58 MG/ML | RxNorm |
| 40166304 | methadone hydrochloride 0.58 MG/ML Oral Solution | RxNorm |
| 40166301 | methadone hydrochloride 0.5 MG/ML | RxNorm |
| 40166302 | methadone hydrochloride 0.5 MG/ML Oral Solution | RxNorm |
| 40166305 | methadone hydrochloride 0.75 MG/ML | RxNorm |
| 40166306 | methadone hydrochloride 0.75 MG/ML Oral Solution | RxNorm |
| 40166319 | methadone hydrochloride 10 MG | RxNorm |
| 40166323 | methadone hydrochloride 10 MG [Dolophine] | RxNorm |
| 40166324 | methadone hydrochloride 10 MG [Methadose] | RxNorm |
| 40166325 | methadone hydrochloride 10 MG/ML | RxNorm |
| 40166328 | methadone hydrochloride 10 MG/ML [Dolophine] | RxNorm |
| 40166326 | methadone hydrochloride 10 MG/ML Injectable Solution | RxNorm |
| 40166327 | methadone hydrochloride 10 MG/ML Injectable Solution [Dolophine] | RxNorm |
| 40220880 | methadone hydrochloride 10 MG/ML [Methadose] | RxNorm |
| 40185005 | methadone hydrochloride 10 MG/ML Oral Solution | RxNorm |
| 40185006 | methadone hydrochloride 10 MG/ML Oral Solution [Methadose] | RxNorm |
| 40166320 | methadone hydrochloride 10 MG Oral Tablet | RxNorm |
| 40166321 | methadone hydrochloride 10 MG Oral Tablet [Dolophine] | RxNorm |
| 40166322 | methadone hydrochloride 10 MG Oral Tablet [Methadose] | RxNorm |
| 40166311 | methadone hydrochloride 1.25 MG/ML | RxNorm |
| 40166312 | methadone hydrochloride 1.25 MG/ML Oral Solution | RxNorm |
| 40166313 | methadone hydrochloride 1.33 MG/ML | RxNorm |
| 40166314 | methadone hydrochloride 1.33 MG/ML Oral Solution | RxNorm |
| 40166315 | methadone hydrochloride 1.5 MG/ML | RxNorm |
| 40166316 | methadone hydrochloride 1.5 MG/ML Oral Solution | RxNorm |
| 40166317 | methadone hydrochloride 1.75 MG/ML | RxNorm |
| 40166318 | methadone hydrochloride 1.75 MG/ML Oral Solution | RxNorm |
| 40166307 | methadone hydrochloride 1 MG | RxNorm |
| 40166309 | methadone hydrochloride 1 MG/ML | RxNorm |
| 40176047 | methadone hydrochloride 1 MG/ML [Martindale Methadone DTF] | RxNorm |
| 40176048 | methadone hydrochloride 1 MG/ML [Methex] | RxNorm |
| 40166310 | methadone hydrochloride 1 MG/ML Oral Solution | RxNorm |
| 19012828 | methadone hydrochloride 1 MG/ML Oral Solution [Martindale Methadone DTF] | RxNorm |
| 19017089 | methadone hydrochloride 1 MG/ML Oral Solution [Methex] | RxNorm |
| 40166308 | methadone hydrochloride 1 MG Oral Tablet | RxNorm |
| 40166339 | methadone hydrochloride 20 MG/ML | RxNorm |
| 40176049 | methadone hydrochloride 20 MG/ML [Methadose] | RxNorm |
| 40166340 | methadone hydrochloride 20 MG/ML Oral Solution | RxNorm |
| 19016305 | methadone hydrochloride 20 MG/ML Oral Solution [Methadose] | RxNorm |
| 40166331 | methadone hydrochloride 2.25 MG/ML | RxNorm |
| 40166332 | methadone hydrochloride 2.25 MG/ML Oral Solution | RxNorm |
| 40166341 | methadone hydrochloride 25 MG | RxNorm |
| 40166333 | methadone hydrochloride 2.5 MG/ML | RxNorm |
| 40166343 | methadone hydrochloride 25 MG/ML | RxNorm |
| 40166344 | methadone hydrochloride 25 MG/ML Injectable Solution | RxNorm |
| 40166334 | methadone hydrochloride 2.5 MG/ML Oral Solution | RxNorm |
| 40166342 | methadone hydrochloride 25 MG Oral Tablet | RxNorm |
| 40166335 | methadone hydrochloride 2.67 MG/ML | RxNorm |
| 40166336 | methadone hydrochloride 2.67 MG/ML Oral Solution | RxNorm |
| 40166337 | methadone hydrochloride 2.75 MG/ML | RxNorm |
| 40166338 | methadone hydrochloride 2.75 MG/ML Oral Solution | RxNorm |
| 40166329 | methadone hydrochloride 2 MG/ML | RxNorm |
| 40166330 | methadone hydrochloride 2 MG/ML Oral Solution | RxNorm |
| 40166347 | methadone hydrochloride 3.25 MG/ML | RxNorm |
| 40166348 | methadone hydrochloride 3.25 MG/ML Oral Solution | RxNorm |
| 40166349 | methadone hydrochloride 3.5 MG/ML | RxNorm |
| 40166350 | methadone hydrochloride 3.5 MG/ML Oral Solution | RxNorm |
| 40166351 | methadone hydrochloride 3.75 MG/ML | RxNorm |
| 40166352 | methadone hydrochloride 3.75 MG/ML Oral Solution | RxNorm |
| 40166345 | methadone hydrochloride 3 MG/ML | RxNorm |
| 40166346 | methadone hydrochloride 3 MG/ML Oral Solution | RxNorm |
| 1594128 | methadone hydrochloride 40 MG | RxNorm |
| 793090 | methadone hydrochloride 40 MG [Diskets] | RxNorm |
| 1594130 | methadone hydrochloride 40 MG [Methadose] | RxNorm |
| 40166295 | methadone hydrochloride 40 MG Tablet for Oral Suspension | RxNorm |
| 793094 | methadone hydrochloride 40 MG Tablet for Oral Suspension [Diskets] | RxNorm |
| 40166296 | methadone hydrochloride 40 MG Tablet for Oral Suspension [Methadose] | RxNorm |
| 40166355 | methadone hydrochloride 4.25 MG/ML | RxNorm |
| 40166356 | methadone hydrochloride 4.25 MG/ML Oral Solution | RxNorm |
| 40166357 | methadone hydrochloride 4.5 MG/ML | RxNorm |
| 40166358 | methadone hydrochloride 4.5 MG/ML Oral Solution | RxNorm |
| 40166359 | methadone hydrochloride 4.75 MG/ML | RxNorm |
| 40166360 | methadone hydrochloride 4.75 MG/ML Oral Solution | RxNorm |
| 40166353 | methadone hydrochloride 4 MG/ML | RxNorm |
| 40166354 | methadone hydrochloride 4 MG/ML Oral Solution | RxNorm |
| 40166369 | methadone hydrochloride 50 MG/ML | RxNorm |
| 40166370 | methadone hydrochloride 50 MG/ML Injectable Solution | RxNorm |
| 40166361 | methadone hydrochloride 5 MG | RxNorm |
| 40166365 | methadone hydrochloride 5 MG [Dolophine] | RxNorm |
| 40166366 | methadone hydrochloride 5 MG [Methadose] | RxNorm |
| 40166367 | methadone hydrochloride 5 MG/ML | RxNorm |
| 40166368 | methadone hydrochloride 5 MG/ML Oral Solution | RxNorm |
| 40166362 | methadone hydrochloride 5 MG Oral Tablet | RxNorm |
| 40166363 | methadone hydrochloride 5 MG Oral Tablet [Dolophine] | RxNorm |
| 40166364 | methadone hydrochloride 5 MG Oral Tablet [Methadose] | RxNorm |
| 19011606 | methadone hydrochloride 5 MG Oral Tablet [Physeptone] | RxNorm |
| 40176050 | methadone hydrochloride 5 MG [Physeptone] | RxNorm |
| 36223152 | methadone Injectable Product | RxNorm |
| 40063779 | methadone Injectable Solution | RxNorm |
| 40064066 | methadone Injectable Solution [Dolophine] | RxNorm |
| 36223153 | methadone Oral Liquid Product | RxNorm |
| 36223154 | methadone Oral Product | RxNorm |
| 40064067 | methadone Oral Solution | RxNorm |
| 40064068 | methadone Oral Solution [Martindale Methadone DTF] | RxNorm |
| 40064069 | methadone Oral Solution [Methadose] | RxNorm |
| 40064070 | methadone Oral Solution [Methex] | RxNorm |
| 40064071 | methadone Oral Tablet | RxNorm |
| 40064072 | methadone Oral Tablet [Dolophine] | RxNorm |
| 40064073 | methadone Oral Tablet [Methadose] | RxNorm |
| 40064074 | methadone Oral Tablet [Physeptone] | RxNorm |
| 36223155 | methadone Pill | RxNorm |
| 1594129 | methadone Tablet for Oral Suspension | RxNorm |
| 793091 | methadone Tablet for Oral Suspension [Diskets] | RxNorm |
| 1594209 | methadone Tablet for Oral Suspension [Methadose] | RxNorm |
| 36239260 | Methadose Oral Liquid Product | RxNorm |
| 36239261 | Methadose Oral Product | RxNorm |
| 36239262 | Methadose Pill | RxNorm |
| 36227849 | Methex Oral Liquid Product | RxNorm |
| 36227850 | Methex Oral Product | RxNorm |
| 36228682 | Physeptone Oral Product | RxNorm |
| 36230019 | Physeptone Pill | RxNorm |
| **Buprenorphine** | | |
| 1201596 | 0.16 ML buprenorphine 50 MG/ML Prefilled Syringe | RxNorm |
| 1201597 | 0.16 ML buprenorphine 50 MG/ML Prefilled Syringe [Brixadi] | RxNorm |
| 1201604 | 0.18 ML buprenorphine 356 MG/ML Prefilled Syringe | RxNorm |
| 1201605 | 0.18 ML buprenorphine 356 MG/ML Prefilled Syringe [Brixadi] | RxNorm |
| 1201606 | 0.27 ML buprenorphine 356 MG/ML Prefilled Syringe | RxNorm |
| 1201607 | 0.27 ML buprenorphine 356 MG/ML Prefilled Syringe [Brixadi] | RxNorm |
| 1201598 | 0.32 ML buprenorphine 50 MG/ML Prefilled Syringe | RxNorm |
| 1201599 | 0.32 ML buprenorphine 50 MG/ML Prefilled Syringe [Brixadi] | RxNorm |
| 1201608 | 0.36 ML buprenorphine 356 MG/ML Prefilled Syringe | RxNorm |
| 1201609 | 0.36 ML buprenorphine 356 MG/ML Prefilled Syringe [Brixadi] | RxNorm |
| 1201600 | 0.48 ML buprenorphine 50 MG/ML Prefilled Syringe | RxNorm |
| 1201601 | 0.48 ML buprenorphine 50 MG/ML Prefilled Syringe [Brixadi] | RxNorm |
| 793477 | 0.5 ML buprenorphine 200 MG/ML Prefilled Syringe | RxNorm |
| 793482 | 0.5 ML buprenorphine 200 MG/ML Prefilled Syringe [Sublocade] | RxNorm |
| 1201602 | 0.64 ML buprenorphine 50 MG/ML Prefilled Syringe | RxNorm |
| 1201603 | 0.64 ML buprenorphine 50 MG/ML Prefilled Syringe [Brixadi] | RxNorm |
| 793569 | 1.5 ML buprenorphine 200 MG/ML Prefilled Syringe | RxNorm |
| 793570 | 1.5 ML buprenorphine 200 MG/ML Prefilled Syringe [Sublocade] | RxNorm |
| 40173407 | 168 HR buprenorphine 0.005 MG/HR Transdermal System | RxNorm |
| 40173408 | 168 HR buprenorphine 0.005 MG/HR Transdermal System [BuTrans] | RxNorm |
| 45776398 | 168 HR buprenorphine 0.0075 MG/HR Transdermal System | RxNorm |
| 45776400 | 168 HR buprenorphine 0.0075 MG/HR Transdermal System [BuTrans] | RxNorm |
| 43559947 | 168 HR buprenorphine 0.015 MG/HR Transdermal System | RxNorm |
| 43559948 | 168 HR buprenorphine 0.015 MG/HR Transdermal System [BuTrans] | RxNorm |
| 40173409 | 168 HR buprenorphine 0.01 MG/HR Transdermal System | RxNorm |
| 40173410 | 168 HR buprenorphine 0.01 MG/HR Transdermal System [BuTrans] | RxNorm |
| 40173411 | 168 HR buprenorphine 0.02 MG/HR Transdermal System | RxNorm |
| 40173412 | 168 HR buprenorphine 0.02 MG/HR Transdermal System [BuTrans] | RxNorm |
| 46234281 | 1 ML buprenorphine 0.3 MG/ML Cartridge | RxNorm |
| 1133223 | 1 ML buprenorphine 0.3 MG/ML Injection | RxNorm |
| 19029477 | 1 ML buprenorphine 0.3 MG/ML Injection [Buprenex] | RxNorm |
| 19101660 | 72 HR buprenorphine 0.035 MG/HR Transdermal System | RxNorm |
| 19101659 | 72 HR buprenorphine 0.0525 MG/HR Transdermal System | RxNorm |
| 19101658 | 72 HR buprenorphine 0.07 MG/HR Transdermal System | RxNorm |
| 36247793 | Animalgesics Injectable Product | RxNorm |
| 36248309 | Belbuca Buccal Product | RxNorm |
| 36248310 | Belbuca Oral Product | RxNorm |
| 1201392 | Brixadi Injectable Product | RxNorm |
| 36247165 | Bunavail Buccal Product | RxNorm |
| 36247166 | Bunavail Oral Product | RxNorm |
| 36233181 | Buprenex Injectable Product | RxNorm |
| 1133201 | buprenorphine | RxNorm |
| 40173763 | buprenorphine 0.005 MG/HR | RxNorm |
| 40173764 | buprenorphine 0.005 MG/HR [BuTrans] | RxNorm |
| 42902413 | buprenorphine 0.005 MG/HR Transdermal System | RxNorm |
| 42903391 | buprenorphine 0.005 MG/HR Transdermal System [BuTrans] | RxNorm |
| 45776397 | buprenorphine 0.0075 MG/HR | RxNorm |
| 45776399 | buprenorphine 0.0075 MG/HR [BuTrans] | RxNorm |
| 45776401 | buprenorphine 0.0075 MG/HR Transdermal System | RxNorm |
| 45776402 | buprenorphine 0.0075 MG/HR Transdermal System [BuTrans] | RxNorm |
| 43560031 | buprenorphine 0.015 MG/HR | RxNorm |
| 43560034 | buprenorphine 0.015 MG/HR [BuTrans] | RxNorm |
| 43560032 | buprenorphine 0.015 MG/HR Transdermal System | RxNorm |
| 43560033 | buprenorphine 0.015 MG/HR Transdermal System [BuTrans] | RxNorm |
| 40173765 | buprenorphine 0.01 MG/HR | RxNorm |
| 40173766 | buprenorphine 0.01 MG/HR [BuTrans] | RxNorm |
| 42902544 | buprenorphine 0.01 MG/HR Transdermal System | RxNorm |
| 42903327 | buprenorphine 0.01 MG/HR Transdermal System [BuTrans] | RxNorm |
| 40173767 | buprenorphine 0.02 MG/HR | RxNorm |
| 40173768 | buprenorphine 0.02 MG/HR [BuTrans] | RxNorm |
| 42903345 | buprenorphine 0.02 MG/HR Transdermal System | RxNorm |
| 42903403 | buprenorphine 0.02 MG/HR Transdermal System [BuTrans] | RxNorm |
| 19102015 | buprenorphine 0.035 MG/HR | RxNorm |
| 42902727 | buprenorphine 0.035 MG/HR Transdermal System | RxNorm |
| 19102016 | buprenorphine 0.0525 MG/HR | RxNorm |
| 42903006 | buprenorphine 0.0525 MG/HR Transdermal System | RxNorm |
| 35602956 | buprenorphine 0.075 MG | RxNorm |
| 35602958 | buprenorphine 0.075 MG [Belbuca] | RxNorm |
| 35602957 | buprenorphine 0.075 MG Buccal Film | RxNorm |
| 35602959 | buprenorphine 0.075 MG Buccal Film [Belbuca] | RxNorm |
| 19102014 | buprenorphine 0.07 MG/HR | RxNorm |
| 42902864 | buprenorphine 0.07 MG/HR Transdermal System | RxNorm |
| 35602937 | buprenorphine 0.15 MG | RxNorm |
| 35602941 | buprenorphine 0.15 MG [Belbuca] | RxNorm |
| 35602939 | buprenorphine 0.15 MG Buccal Film | RxNorm |
| 35602943 | buprenorphine 0.15 MG Buccal Film [Belbuca] | RxNorm |
| 1133227 | buprenorphine 0.2 MG | RxNorm |
| 1133224 | buprenorphine 0.2 MG Sublingual Tablet | RxNorm |
| 19006098 | buprenorphine 0.2 MG Sublingual Tablet [Subutex] | RxNorm |
| 19116020 | buprenorphine 0.2 MG [Subutex] | RxNorm |
| 35602944 | buprenorphine 0.3 MG | RxNorm |
| 35602946 | buprenorphine 0.3 MG [Belbuca] | RxNorm |
| 35602945 | buprenorphine 0.3 MG Buccal Film | RxNorm |
| 35602947 | buprenorphine 0.3 MG Buccal Film [Belbuca] | RxNorm |
| 19086196 | buprenorphine 0.3 MG/ML | RxNorm |
| 19048353 | buprenorphine 0.3 MG/ML [Buprenex] | RxNorm |
| 46234282 | buprenorphine 0.3 MG/ML Cartridge | RxNorm |
| 35605174 | buprenorphine 0.3 MG/ML Injection | RxNorm |
| 35605175 | buprenorphine 0.3 MG/ML Injection [Buprenex] | RxNorm |
| 35602948 | buprenorphine 0.45 MG | RxNorm |
| 35602950 | buprenorphine 0.45 MG [Belbuca] | RxNorm |
| 35602949 | buprenorphine 0.45 MG Buccal Film | RxNorm |
| 35602951 | buprenorphine 0.45 MG Buccal Film [Belbuca] | RxNorm |
| 1133228 | buprenorphine 0.4 MG | RxNorm |
| 19062756 | buprenorphine 0.4 MG Sublingual Tablet | RxNorm |
| 19006099 | buprenorphine 0.4 MG Sublingual Tablet [Subutex] | RxNorm |
| 19116021 | buprenorphine 0.4 MG [Subutex] | RxNorm |
| 35602952 | buprenorphine 0.6 MG | RxNorm |
| 35602954 | buprenorphine 0.6 MG [Belbuca] | RxNorm |
| 35602953 | buprenorphine 0.6 MG Buccal Film | RxNorm |
| 35602955 | buprenorphine 0.6 MG Buccal Film [Belbuca] | RxNorm |
| 35602960 | buprenorphine 0.75 MG | RxNorm |
| 35602962 | buprenorphine 0.75 MG [Belbuca] | RxNorm |
| 35602961 | buprenorphine 0.75 MG Buccal Film | RxNorm |
| 35602963 | buprenorphine 0.75 MG Buccal Film [Belbuca] | RxNorm |
| 1593216 | buprenorphine 0.7 MG | RxNorm |
| 1593218 | buprenorphine 0.7 MG / naloxone 0.18 MG Sublingual Tablet | RxNorm |
| 1593220 | buprenorphine 0.7 MG / naloxone 0.18 MG Sublingual Tablet [Zubsolv] | RxNorm |
| 1593219 | buprenorphine 0.7 MG / naloxone 0.18 MG [Zubsolv] | RxNorm |
| 35602964 | buprenorphine 0.9 MG | RxNorm |
| 35602966 | buprenorphine 0.9 MG [Belbuca] | RxNorm |
| 35602965 | buprenorphine 0.9 MG Buccal Film | RxNorm |
| 35602967 | buprenorphine 0.9 MG Buccal Film [Belbuca] | RxNorm |
| 45892565 | buprenorphine 11.4 MG | RxNorm |
| 45892567 | buprenorphine 11.4 MG / naloxone 2.9 MG Sublingual Tablet | RxNorm |
| 45892569 | buprenorphine 11.4 MG / naloxone 2.9 MG Sublingual Tablet [Zubsolv] | RxNorm |
| 45892568 | buprenorphine 11.4 MG / naloxone 2.9 MG [Zubsolv] | RxNorm |
| 42898498 | buprenorphine 12 MG | RxNorm |
| 42898499 | buprenorphine 12 MG / naloxone 3 MG Sublingual Film | RxNorm |
| 42898500 | buprenorphine 12 MG / naloxone 3 MG Sublingual Film [Suboxone] | RxNorm |
| 42898501 | buprenorphine 12 MG / naloxone 3 MG [Suboxone] | RxNorm |
| 43532742 | buprenorphine 1.4 MG | RxNorm |
| 43532942 | buprenorphine 1.4 MG / naloxone 0.36 MG Sublingual Tablet | RxNorm |
| 43532943 | buprenorphine 1.4 MG / naloxone 0.36 MG Sublingual Tablet [Zubsolv] | RxNorm |
| 43532745 | buprenorphine 1.4 MG / naloxone 0.36 MG [Zubsolv] | RxNorm |
| 35200724 | buprenorphine 16 MG | RxNorm |
| 1355907 | buprenorphine 16 MG / naloxone 4 MG [Cassipa] | RxNorm |
| 35200725 | buprenorphine 16 MG / naloxone 4 MG Sublingual Film | RxNorm |
| 1355911 | buprenorphine 16 MG / naloxone 4 MG Sublingual Film [Cassipa] | RxNorm |
| 45776066 | buprenorphine 1.8 MG/ML | RxNorm |
| 45776067 | buprenorphine 1.8 MG/ML Injectable Solution | RxNorm |
| 45776190 | buprenorphine 1.8 MG/ML Injectable Solution [Simbadol] | RxNorm |
| 45776188 | buprenorphine 1.8 MG/ML [Simbadol] | RxNorm |
| 793475 | buprenorphine 200 MG/ML | RxNorm |
| 793567 | buprenorphine 200 MG/ML Prefilled Syringe | RxNorm |
| 793568 | buprenorphine 200 MG/ML Prefilled Syringe [Sublocade] | RxNorm |
| 793479 | buprenorphine 200 MG/ML [Sublocade] | RxNorm |
| 779546 | buprenorphine 20 MG/ML | RxNorm |
| 779548 | buprenorphine 20 MG/ML Topical Solution | RxNorm |
| 779553 | buprenorphine 20 MG/ML Topical Solution [Zorbium] | RxNorm |
| 779550 | buprenorphine 20 MG/ML [Zorbium] | RxNorm |
| 45776268 | buprenorphine 2.1 MG | RxNorm |
| 45776271 | buprenorphine 2.1 MG / naloxone 0.3 MG Buccal Film | RxNorm |
| 45776275 | buprenorphine 2.1 MG / naloxone 0.3 MG Buccal Film [Bunavail] | RxNorm |
| 45776273 | buprenorphine 2.1 MG / naloxone 0.3 MG [Bunavail] | RxNorm |
| 46287541 | buprenorphine 2.9 MG | RxNorm |
| 46287543 | buprenorphine 2.9 MG / naloxone 0.71 MG Sublingual Tablet | RxNorm |
| 46287551 | buprenorphine 2.9 MG / naloxone 0.71 MG Sublingual Tablet [Zubsolv] | RxNorm |
| 46287550 | buprenorphine 2.9 MG / naloxone 0.71 MG [Zubsolv] | RxNorm |
| 19098731 | buprenorphine 2 MG | RxNorm |
| 40225989 | buprenorphine 2 MG / naloxone 0.5 MG Sublingual Film | RxNorm |
| 40225990 | buprenorphine 2 MG / naloxone 0.5 MG Sublingual Film [Suboxone] | RxNorm |
| 1133231 | buprenorphine 2 MG / naloxone 0.5 MG Sublingual Tablet | RxNorm |
| 40225991 | buprenorphine 2 MG / naloxone 0.5 MG Sublingual Tablet [Suboxone] | RxNorm |
| 40225992 | buprenorphine 2 MG / naloxone 0.5 MG [Suboxone] | RxNorm |
| 1133229 | buprenorphine 2 MG Sublingual Tablet | RxNorm |
| 19102738 | buprenorphine 2 MG Sublingual Tablet [Subutex] | RxNorm |
| 19033208 | buprenorphine 2 MG [Subutex] | RxNorm |
| 1201395 | buprenorphine 356 MG/ML | RxNorm |
| 1201397 | buprenorphine 356 MG/ML [Brixadi] | RxNorm |
| 1201396 | buprenorphine 356 MG/ML Prefilled Syringe | RxNorm |
| 1201398 | buprenorphine 356 MG/ML Prefilled Syringe [Brixadi] | RxNorm |
| 45774516 | buprenorphine 4.2 MG | RxNorm |
| 45774518 | buprenorphine 4.2 MG / naloxone 0.7 MG Buccal Film | RxNorm |
| 45774520 | buprenorphine 4.2 MG / naloxone 0.7 MG Buccal Film [Bunavail] | RxNorm |
| 45774519 | buprenorphine 4.2 MG / naloxone 0.7 MG [Bunavail] | RxNorm |
| 42898502 | buprenorphine 4 MG | RxNorm |
| 42898503 | buprenorphine 4 MG / naloxone 1 MG Sublingual Film | RxNorm |
| 42898504 | buprenorphine 4 MG / naloxone 1 MG Sublingual Film [Suboxone] | RxNorm |
| 42898505 | buprenorphine 4 MG / naloxone 1 MG [Suboxone] | RxNorm |
| 1201388 | buprenorphine 50 MG/ML | RxNorm |
| 1201391 | buprenorphine 50 MG/ML [Brixadi] | RxNorm |
| 1201389 | buprenorphine 50 MG/ML Prefilled Syringe | RxNorm |
| 1201394 | buprenorphine 50 MG/ML Prefilled Syringe [Brixadi] | RxNorm |
| 43532747 | buprenorphine 5.7 MG | RxNorm |
| 43532944 | buprenorphine 5.7 MG / naloxone 1.4 MG Sublingual Tablet | RxNorm |
| 43532945 | buprenorphine 5.7 MG / naloxone 1.4 MG Sublingual Tablet [Zubsolv] | RxNorm |
| 43532749 | buprenorphine 5.7 MG / naloxone 1.4 MG [Zubsolv] | RxNorm |
| 45776279 | buprenorphine 6.3 MG | RxNorm |
| 45774521 | buprenorphine 6.3 MG / naloxone 1 MG Buccal Film | RxNorm |
| 45774523 | buprenorphine 6.3 MG / naloxone 1 MG Buccal Film [Bunavail] | RxNorm |
| 45774522 | buprenorphine 6.3 MG / naloxone 1 MG [Bunavail] | RxNorm |
| 36250047 | buprenorphine 74.2 MG | RxNorm |
| 36250050 | buprenorphine 74.2 MG Drug Implant | RxNorm |
| 36250063 | buprenorphine 74.2 MG Drug Implant [Probuphine] | RxNorm |
| 36250052 | buprenorphine 74.2 MG [Probuphine] | RxNorm |
| 45892570 | buprenorphine 8.6 MG | RxNorm |
| 45892572 | buprenorphine 8.6 MG / naloxone 2.1 MG Sublingual Tablet | RxNorm |
| 45892574 | buprenorphine 8.6 MG / naloxone 2.1 MG Sublingual Tablet [Zubsolv] | RxNorm |
| 45892573 | buprenorphine 8.6 MG / naloxone 2.1 MG [Zubsolv] | RxNorm |
| 1133266 | buprenorphine 8 MG | RxNorm |
| 40225993 | buprenorphine 8 MG / naloxone 2 MG Sublingual Film | RxNorm |
| 40225994 | buprenorphine 8 MG / naloxone 2 MG Sublingual Film [Suboxone] | RxNorm |
| 1133262 | buprenorphine 8 MG / naloxone 2 MG Sublingual Tablet | RxNorm |
| 40225995 | buprenorphine 8 MG / naloxone 2 MG Sublingual Tablet [Suboxone] | RxNorm |
| 40225996 | buprenorphine 8 MG / naloxone 2 MG [Suboxone] | RxNorm |
| 1133230 | buprenorphine 8 MG Sublingual Tablet | RxNorm |
| 19102739 | buprenorphine 8 MG Sublingual Tablet [Subutex] | RxNorm |
| 19033209 | buprenorphine 8 MG [Subutex] | RxNorm |
| 35602938 | buprenorphine Buccal Film | RxNorm |
| 35602942 | buprenorphine Buccal Film [Belbuca] | RxNorm |
| 36248308 | buprenorphine Buccal Product | RxNorm |
| 46234280 | buprenorphine Cartridge | RxNorm |
| 36250049 | buprenorphine Drug Implant | RxNorm |
| 36250053 | buprenorphine Drug Implant [Probuphine] | RxNorm |
| 36250048 | buprenorphine Drug Implant Product | RxNorm |
| 44785597 | buprenorphine hydrochloride 1.3 MG/ML | RxNorm |
| 44785600 | buprenorphine hydrochloride 1.3 MG/ML [Animalgesics] | RxNorm |
| 44785598 | buprenorphine hydrochloride 1.3 MG/ML Injectable Suspension | RxNorm |
| 44785599 | buprenorphine hydrochloride 1.3 MG/ML Injectable Suspension [Animalgesics] | RxNorm |
| 36217860 | buprenorphine Injectable Product | RxNorm |
| 40015178 | buprenorphine Injectable Solution | RxNorm |
| 45776189 | buprenorphine Injectable Solution [Simbadol] | RxNorm |
| 44785595 | buprenorphine Injectable Suspension | RxNorm |
| 44785596 | buprenorphine Injectable Suspension [Animalgesics] | RxNorm |
| 35605172 | buprenorphine Injection | RxNorm |
| 35605173 | buprenorphine Injection [Buprenex] | RxNorm |
| 45776270 | buprenorphine / naloxone Buccal Film | RxNorm |
| 45776274 | buprenorphine / naloxone Buccal Film [Bunavail] | RxNorm |
| 36247164 | buprenorphine / naloxone Buccal Product | RxNorm |
| 37498349 | buprenorphine / naloxone Oral Film Product | RxNorm |
| 36217858 | buprenorphine / naloxone Oral Product | RxNorm |
| 36217859 | buprenorphine / naloxone Pill | RxNorm |
| 37498350 | buprenorphine / naloxone Sublingual Film | RxNorm |
| 37498388 | buprenorphine / naloxone Sublingual Film [Cassipa] | RxNorm |
| 37498351 | buprenorphine / naloxone Sublingual Film [Suboxone] | RxNorm |
| 36244326 | buprenorphine / naloxone Sublingual Product | RxNorm |
| 40015149 | buprenorphine / naloxone Sublingual Tablet | RxNorm |
| 40225988 | buprenorphine / naloxone Sublingual Tablet [Suboxone] | RxNorm |
| 43532746 | buprenorphine / naloxone Sublingual Tablet [Zubsolv] | RxNorm |
| 36217861 | buprenorphine Oral Product | RxNorm |
| 36217862 | buprenorphine Pill | RxNorm |
| 793476 | buprenorphine Prefilled Syringe | RxNorm |
| 1201393 | buprenorphine Prefilled Syringe [Brixadi] | RxNorm |
| 793480 | buprenorphine Prefilled Syringe [Sublocade] | RxNorm |
| 36244872 | buprenorphine Sublingual Product | RxNorm |
| 40015181 | buprenorphine Sublingual Tablet | RxNorm |
| 40015182 | buprenorphine Sublingual Tablet [Subutex] | RxNorm |
| 36217863 | buprenorphine Topical Product | RxNorm |
| 779547 | buprenorphine Topical Solution | RxNorm |
| 779552 | buprenorphine Topical Solution [Zorbium] | RxNorm |
| 36245276 | buprenorphine Transdermal Product | RxNorm |
| 40221142 | buprenorphine Transdermal System | RxNorm |
| 40221143 | buprenorphine Transdermal System [BuTrans] | RxNorm |
| 36219772 | BuTrans Topical Product | RxNorm |
| 36245397 | BuTrans Transdermal Product | RxNorm |
| 37498375 | Cassipa Oral Film Product | RxNorm |
| 1355909 | Cassipa Oral Product | RxNorm |
| 37498389 | Cassipa Sublingual Product | RxNorm |
| 36250054 | Probuphine Drug Implant Product | RxNorm |
| 36248720 | Simbadol Injectable Product | RxNorm |
| 793481 | Sublocade Injectable Product | RxNorm |
| 37498352 | Suboxone Oral Film Product | RxNorm |
| 36232564 | Suboxone Oral Product | RxNorm |
| 36232565 | Suboxone Pill | RxNorm |
| 36244939 | Suboxone Sublingual Product | RxNorm |
| 36232566 | Subutex Oral Product | RxNorm |
| 36232567 | Subutex Pill | RxNorm |
| 36243968 | Subutex Sublingual Product | RxNorm |
| 779551 | Zorbium Topical Product | RxNorm |
| 36248031 | Zubsolv Oral Product | RxNorm |
| 36248032 | Zubsolv Pill | RxNorm |
| 36248033 | Zubsolv Sublingual Product | RxNorm |
| **Naltrexone** | | |
| 45774486 | 12 HR bupropion hydrochloride 90 MG / naltrexone hydrochloride 8 MG Extended Release Oral Tablet | RxNorm |
| 45774490 | 12 HR bupropion hydrochloride 90 MG / naltrexone hydrochloride 8 MG Extended Release Oral Tablet [Contrave] | RxNorm |
| 40221196 | 12 HR naltrexone hydrochloride 1.2 MG / oxycodone hydrochloride 10 MG Extended Release Oral Capsule | RxNorm |
| 40221202 | 12 HR naltrexone hydrochloride 1.2 MG / oxycodone hydrochloride 10 MG Extended Release Oral Capsule [Troxyca] | RxNorm |
| 40221205 | 12 HR naltrexone hydrochloride 2.4 MG / oxycodone hydrochloride 20 MG Extended Release Oral Capsule | RxNorm |
| 40221207 | 12 HR naltrexone hydrochloride 2.4 MG / oxycodone hydrochloride 20 MG Extended Release Oral Capsule [Troxyca] | RxNorm |
| 40221211 | 12 HR naltrexone hydrochloride 3.6 MG / oxycodone hydrochloride 30 MG Extended Release Oral Capsule | RxNorm |
| 40221213 | 12 HR naltrexone hydrochloride 3.6 MG / oxycodone hydrochloride 30 MG Extended Release Oral Capsule [Troxyca] | RxNorm |
| 40221217 | 12 HR naltrexone hydrochloride 4.8 MG / oxycodone hydrochloride 40 MG Extended Release Oral Capsule | RxNorm |
| 40221219 | 12 HR naltrexone hydrochloride 4.8 MG / oxycodone hydrochloride 40 MG Extended Release Oral Capsule [Troxyca] | RxNorm |
| 40221223 | 12 HR naltrexone hydrochloride 7.2 MG / oxycodone hydrochloride 60 MG Extended Release Oral Capsule | RxNorm |
| 40221225 | 12 HR naltrexone hydrochloride 7.2 MG / oxycodone hydrochloride 60 MG Extended Release Oral Capsule [Troxyca] | RxNorm |
| 40221229 | 12 HR naltrexone hydrochloride 9.6 MG / oxycodone hydrochloride 80 MG Extended Release Oral Capsule | RxNorm |
| 40221231 | 12 HR naltrexone hydrochloride 9.6 MG / oxycodone hydrochloride 80 MG Extended Release Oral Capsule [Troxyca] | RxNorm |
| 40166392 | Abuse-Deterrent morphine sulfate 100 MG / naltrexone hydrochloride 4 MG Extended Release Oral Capsule | RxNorm |
| 40166393 | Abuse-Deterrent morphine sulfate 100 MG / naltrexone hydrochloride 4 MG Extended Release Oral Capsule [Embeda] | RxNorm |
| 40166395 | Abuse-Deterrent morphine sulfate 20 MG / naltrexone hydrochloride 0.8 MG Extended Release Oral Capsule | RxNorm |
| 40166396 | Abuse-Deterrent morphine sulfate 20 MG / naltrexone hydrochloride 0.8 MG Extended Release Oral Capsule [Embeda] | RxNorm |
| 40166398 | Abuse-Deterrent morphine sulfate 30 MG / naltrexone hydrochloride 1.2 MG Extended Release Oral Capsule | RxNorm |
| 40166399 | Abuse-Deterrent morphine sulfate 30 MG / naltrexone hydrochloride 1.2 MG Extended Release Oral Capsule [Embeda] | RxNorm |
| 40166400 | Abuse-Deterrent morphine sulfate 50 MG / naltrexone hydrochloride 2 MG Extended Release Oral Capsule | RxNorm |
| 40166401 | Abuse-Deterrent morphine sulfate 50 MG / naltrexone hydrochloride 2 MG Extended Release Oral Capsule [Embeda] | RxNorm |
| 40166403 | Abuse-Deterrent morphine sulfate 60 MG / naltrexone hydrochloride 2.4 MG Extended Release Oral Capsule | RxNorm |
| 40166404 | Abuse-Deterrent morphine sulfate 60 MG / naltrexone hydrochloride 2.4 MG Extended Release Oral Capsule [Embeda] | RxNorm |
| 40166407 | Abuse-Deterrent morphine sulfate 80 MG / naltrexone hydrochloride 3.2 MG Extended Release Oral Capsule | RxNorm |
| 40166408 | Abuse-Deterrent morphine sulfate 80 MG / naltrexone hydrochloride 3.2 MG Extended Release Oral Capsule [Embeda] | RxNorm |
| 45774488 | bupropion hydrochloride 90 MG / naltrexone hydrochloride 8 MG [Contrave] | RxNorm |
| 45774491 | bupropion hydrochloride 90 MG / naltrexone hydrochloride 8 MG Extended Release Oral Tablet | RxNorm |
| 45774492 | bupropion hydrochloride 90 MG / naltrexone hydrochloride 8 MG Extended Release Oral Tablet [Contrave] | RxNorm |
| 45774485 | bupropion / naltrexone Extended Release Oral Tablet | RxNorm |
| 45774489 | bupropion / naltrexone Extended Release Oral Tablet [Contrave] | RxNorm |
| 36249045 | bupropion / naltrexone Oral Product | RxNorm |
| 36249046 | bupropion / naltrexone Pill | RxNorm |
| 36249047 | Contrave Oral Product | RxNorm |
| 36249048 | Contrave Pill | RxNorm |
| 36237170 | Depade Oral Product | RxNorm |
| 36237171 | Depade Pill | RxNorm |
| 36228117 | Embeda Oral Product | RxNorm |
| 36228118 | Embeda Pill | RxNorm |
| 40166388 | morphine / naltrexone Extended Release Oral Capsule | RxNorm |
| 40166389 | morphine / naltrexone Extended Release Oral Capsule [Embeda] | RxNorm |
| 36219425 | morphine / naltrexone Oral Product | RxNorm |
| 36219426 | morphine / naltrexone Pill | RxNorm |
| 40166394 | morphine sulfate 100 MG / naltrexone hydrochloride 4 MG [Embeda] | RxNorm |
| 40165049 | morphine sulfate 20 MG / naltrexone hydrochloride 0.8 MG [Embeda] | RxNorm |
| 40165053 | morphine sulfate 30 MG / naltrexone hydrochloride 1.2 MG [Embeda] | RxNorm |
| 40165057 | morphine sulfate 50 MG / naltrexone hydrochloride 2 MG [Embeda] | RxNorm |
| 40166405 | morphine sulfate 60 MG / naltrexone hydrochloride 2.4 MG [Embeda] | RxNorm |
| 40166409 | morphine sulfate 80 MG / naltrexone hydrochloride 3.2 MG [Embeda] | RxNorm |
| 1714319 | naltrexone | RxNorm |
| 35605492 | naltrexone 380 MG | RxNorm |
| 1714351 | naltrexone 380 MG Injection | RxNorm |
| 1714372 | naltrexone 380 MG Injection [Vivitrol] | RxNorm |
| 35605494 | naltrexone 380 MG [Vivitrol] | RxNorm |
| 40165108 | naltrexone hydrochloride 0.8 MG | RxNorm |
| 44784869 | naltrexone hydrochloride 100 MG | RxNorm |
| 44784872 | naltrexone hydrochloride 100 MG [Depade] | RxNorm |
| 44784870 | naltrexone hydrochloride 100 MG Oral Tablet | RxNorm |
| 44784871 | naltrexone hydrochloride 100 MG Oral Tablet [Depade] | RxNorm |
| 40165109 | naltrexone hydrochloride 1.2 MG | RxNorm |
| 40221203 | naltrexone hydrochloride 1.2 MG / oxycodone hydrochloride 10 MG Extended Release Oral Capsule | RxNorm |
| 40221204 | naltrexone hydrochloride 1.2 MG / oxycodone hydrochloride 10 MG Extended Release Oral Capsule [Troxyca] | RxNorm |
| 40221198 | naltrexone hydrochloride 1.2 MG / oxycodone hydrochloride 10 MG [Troxyca] | RxNorm |
| 40166413 | naltrexone hydrochloride 2.4 MG | RxNorm |
| 40221208 | naltrexone hydrochloride 2.4 MG / oxycodone hydrochloride 20 MG Extended Release Oral Capsule | RxNorm |
| 40221209 | naltrexone hydrochloride 2.4 MG / oxycodone hydrochloride 20 MG Extended Release Oral Capsule [Troxyca] | RxNorm |
| 40221206 | naltrexone hydrochloride 2.4 MG / oxycodone hydrochloride 20 MG [Troxyca] | RxNorm |
| 44784873 | naltrexone hydrochloride 25 MG | RxNorm |
| 44784876 | naltrexone hydrochloride 25 MG [Depade] | RxNorm |
| 44784874 | naltrexone hydrochloride 25 MG Oral Tablet | RxNorm |
| 44784875 | naltrexone hydrochloride 25 MG Oral Tablet [Depade] | RxNorm |
| 40165110 | naltrexone hydrochloride 2 MG | RxNorm |
| 40166414 | naltrexone hydrochloride 3.2 MG | RxNorm |
| 40221210 | naltrexone hydrochloride 3.6 MG | RxNorm |
| 40221214 | naltrexone hydrochloride 3.6 MG / oxycodone hydrochloride 30 MG Extended Release Oral Capsule | RxNorm |
| 40221215 | naltrexone hydrochloride 3.6 MG / oxycodone hydrochloride 30 MG Extended Release Oral Capsule [Troxyca] | RxNorm |
| 40221212 | naltrexone hydrochloride 3.6 MG / oxycodone hydrochloride 30 MG [Troxyca] | RxNorm |
| 40221216 | naltrexone hydrochloride 4.8 MG | RxNorm |
| 40221220 | naltrexone hydrochloride 4.8 MG / oxycodone hydrochloride 40 MG Extended Release Oral Capsule | RxNorm |
| 40221221 | naltrexone hydrochloride 4.8 MG / oxycodone hydrochloride 40 MG Extended Release Oral Capsule [Troxyca] | RxNorm |
| 40221218 | naltrexone hydrochloride 4.8 MG / oxycodone hydrochloride 40 MG [Troxyca] | RxNorm |
| 40166415 | naltrexone hydrochloride 4 MG | RxNorm |
| 44784877 | naltrexone hydrochloride 50 MG | RxNorm |
| 44784881 | naltrexone hydrochloride 50 MG [Depade] | RxNorm |
| 44784878 | naltrexone hydrochloride 50 MG Oral Tablet | RxNorm |
| 44784879 | naltrexone hydrochloride 50 MG Oral Tablet [Depade] | RxNorm |
| 44784880 | naltrexone hydrochloride 50 MG Oral Tablet [ReVia] | RxNorm |
| 44784882 | naltrexone hydrochloride 50 MG [ReVia] | RxNorm |
| 40221222 | naltrexone hydrochloride 7.2 MG | RxNorm |
| 40221226 | naltrexone hydrochloride 7.2 MG / oxycodone hydrochloride 60 MG Extended Release Oral Capsule | RxNorm |
| 40221227 | naltrexone hydrochloride 7.2 MG / oxycodone hydrochloride 60 MG Extended Release Oral Capsule [Troxyca] | RxNorm |
| 40221224 | naltrexone hydrochloride 7.2 MG / oxycodone hydrochloride 60 MG [Troxyca] | RxNorm |
| 45774484 | naltrexone hydrochloride 8 MG | RxNorm |
| 40221228 | naltrexone hydrochloride 9.6 MG | RxNorm |
| 40221232 | naltrexone hydrochloride 9.6 MG / oxycodone hydrochloride 80 MG Extended Release Oral Capsule | RxNorm |
| 40221233 | naltrexone hydrochloride 9.6 MG / oxycodone hydrochloride 80 MG Extended Release Oral Capsule [Troxyca] | RxNorm |
| 40221230 | naltrexone hydrochloride 9.6 MG / oxycodone hydrochloride 80 MG [Troxyca] | RxNorm |
| 36212591 | naltrexone Injectable Product | RxNorm |
| 35605493 | naltrexone Injection | RxNorm |
| 35605495 | naltrexone Injection [Vivitrol] | RxNorm |
| 36212592 | naltrexone Oral Product | RxNorm |
| 40063517 | naltrexone Oral Tablet | RxNorm |
| 40117295 | naltrexone Oral Tablet [Depade] | RxNorm |
| 40063518 | naltrexone Oral Tablet [ReVia] | RxNorm |
| 40221195 | naltrexone / oxycodone Extended Release Oral Capsule | RxNorm |
| 40221199 | naltrexone / oxycodone Extended Release Oral Capsule [Troxyca] | RxNorm |
| 40221193 | naltrexone / oxycodone Oral Product | RxNorm |
| 40221194 | naltrexone / oxycodone Pill | RxNorm |
| 36212593 | naltrexone Pill | RxNorm |
| 36232090 | ReVia Oral Product | RxNorm |
| 36232091 | ReVia Pill | RxNorm |
| 40221200 | Troxyca Oral Product | RxNorm |
| 40221201 | Troxyca Pill | RxNorm |
| 36239345 | Vivitrol Injectable Product | RxNorm |
